# Longitudinal MRI reveals adolescent pituitary growth patterns linked to puberty and environmental exposures

**DOI:** 10.64898/2026.06.26.26356701

**Authors:** Giorgia Picci, Deanna M. Barch, Bart Larsen, Carina Heller, Damien A. Fair, Bianca T. Leonard, Leah H. Somerville, Michael A. Yassa, Hal S. Stern, Lynsie R. Warr, Elizabeth A. Shirtcliff, Jerod M. Rasmussen

## Abstract

Puberty is a period of profound behavioral reorganization that recalibrates social motivation, risk-taking, and sexual behavior in ways that shape lifelong human health. Yet its characterization in population-based studies relies largely on self-report, which reflects perceived physical changes rather than the neuroendocrine substrates driving the transition. The pituitary gland sits at the center of this reorganization, coordinating hypothalamic-pituitary axes that orchestrate puberty. Leveraging 11,818 adolescents (ABCD Study; 30,276 MRI observations), we show that pituitary volume is a precise, scalable marker of pubertal progression carrying non-redundant information beyond chronological age, salivary hormones, and self-reported stage. Sex-specific non-linear trajectories, accelerated expansion anchored to menarche, and distinct patterns across menarche timing subgroups capture both the timing and tempo of puberty at population scale. Pituitary volume was further associated with ACEs and decreased within-person growth following hormonal contraception initiation, positioning it as a sensitive index of the biological embedding of exogenous exposures known to influence pubertal maturation.

## 1. Introduction

Puberty reorganizes human behavior perhaps more profoundly than any other developmental transition, recalibrating social motivation, risk-taking, reward sensitivity, and sexual behavior in ways that shape mental health and adaptive functioning across the lifespan. Its hormonal surges and neurodevelopmental changes initiate cascading effects on brain structure (1), social cognition, and emotional regulation (2). Individual differences in pubertal timing and tempo are among the most consistent predictors of internalizing and externalizing symptoms (3–5), substance use (6, 7), and altered social relationships (2), with effects that often persist into adulthood (8, 9). This sensitivity reflects a coupling between biological maturation and social context (i.e., developmental windows (10, 11)), in which lived experience becomes biologically embedded, shaping vulnerability. Identifying who is progressing through this window, and at what pace, therefore requires biological markers that track maturation itself rather than its downstream outputs.

This maturational process is driven by a major reawakening and remodeling of neuroendocrine systems, orchestrated by coordinated activity across multiple hypothalamic-pituitary axes. These axes regulate somatic growth, reproductive maturation, stress physiology, and metabolism. Central to these processes is the pituitary gland, comprised of two anatomically and functionally distinct lobes (*i.e.,* anterior and posterior) located just inferior to the hypothalamus. The anterior pituitary primarily synthesizes and releases trophic hormones (*e.g.,* luteinizing hormone, follicle-stimulating hormone, and adrenocorticotropic hormone) that regulate downstream production of gonadal and adrenal steroids such as estradiol, testosterone within the hypothalamic-pituitary-gonadal (HPG, reproductive maturation) axes and cortisol within the hypothalamic-pituitary-adrenal (HPA, stress physiology), as well as other developmentally-salient hormones like prolactin and growth hormones (12). In contrast, the posterior pituitary is responsible for the storage and release of vasopressin and oxytocin peptides (13), and does not initiate or regulate endocrine cascades that underlie pubertal maturation. Accordingly, the anterior pituitary plays a central role in pubertal and stress-related neuroendocrine signaling, acting as a transducer that integrates hypothalamic inputs into systemic endocrine cascades that governs the timing and tempo of maturation.

MRI-based studies have been instrumental in demonstrating that pituitary morphology tracks circulating pubertal hormone levels (14–17) and physical indicators of puberty (18, 19), and have demonstrated plasticity in the context of environmental exposures (*e.g.,* adversity) (20–23). Structural MRI studies demonstrate that pituitary volume increases throughout childhood and adolescence, with females exhibiting larger volumes and an earlier onset of puberty-related expansion (16, 24) that parallels known sex differences in pubertal timing. Accelerated pituitary growth following early-life psychosocial stress has also been observed (20, 23, 25), with enlargement persisting into adulthood (26). Importantly, emerging work has demonstrated that MRI-derived pituitary volume is sensitive enough to index rapid within-person changes in endocrine milieu during time-limited periods of hormonal flux (*e.g.,* pregnancy) (27). While these studies support the utility of in vivo characterization of the pituitary as a window into endocrine capacity, recent evidence suggests that traditional-scale investigations are underpowered to reliably estimate the more nuanced individual differences and non-linear dynamics that tend to be a hallmark of the puberty transition (28). In addition, an overwhelming focus on the whole pituitary gland rather than its two distinct lobes, and narrow temporal coverage of the pubertal process and its contextual influences, hinders our ability to understand how environmental experiences are biologically embedded in pituitary development to shape the life course. Recent advances in automated machine learning-based segmentation now offer the opportunity to overcome longstanding technical barriers, enabling scalable, longitudinal characterization of pituitary development.

Automated tools for pituitary segmentation have begun to emerge (29, 30) (31, 32), including a recent 3D U-net implementation within FreeSurfer (33). However, these models have been trained on stable adult anatomy and thus may not fully capture the rapid morphological changes and signal variations characteristic of the maturing adolescent brain and pituitary region, which also make registration-based methods challenging (*e.g.,* pneumatization status of the sphenoid sinus (34)). To address this, and be consistent with other recent neural-network based approaches (29, 33), we developed a study-specific two-stage 3D U-net pipeline optimized for high-throughput segmentation in pediatric and adolescent samples. Leveraging these recent computational advancements and the unprecedented scale of the Adolescent Brain Cognitive Development (ABCD) study, we applied this framework to over 30,000 MRI scans from more than 11,000 youth, enabling the first population-level mapping of pituitary development across the pubertal transition.

The present study builds upon this normative framework and the ABCD study’s rich phenotyping of pubertal processes (*e.g.,* repeated MRI assessments, self-reported pubertal status, salivary hormones, environmental measures) to test hypotheses centered on sex differences and interindividual variability around developmental trajectories. Specifically, we evaluated the utility of anterior pituitary volume as a structural surrogate for pubertal tempo and age at menarche, testing whether this image-based phenotype provides unique information regarding maturation beyond circulating pubertal hormone levels alone. Furthermore, we assessed the plasticity of pituitary development in response to external modulation, examining its sensitivity to both adverse childhood experiences and the introduction of exogenous hormonal contraceptives. To ensure the rigor of these observations, we established measurement validity by cross-validating our findings against the newly available FreeSurfer pituitary pipeline (33) and demonstrated biological validity by confirming that developmental effects were consistently driven by the functionally dominant anterior lobe. By integrating these population-scale dynamics with clinical and environmental markers, we position the pituitary as a central, measurable landmark of the neuroendocrine transition and a sensitive index of the biological embedding of experience known to influence pubertal maturation (24).

## 2. Results

### 2.1 Automated pituitary segmentation and validation

An ABCD-specific two-stage 3D U-Net pipeline was used to segment anterior and posterior pituitary lobes across 30,276 T1-weighted scans from 11,818 ABCD participants, with total end-to-end inference time of <3 s per scan. A manually labeled test set was used for development and tracking performance across training epochs (n = 14), and performance was quantified using a measure of spatial overlap between inferred and ground truth labels (Dice Similarity Coefficient). Observed performance supported successful stage one bounding box localization in low-resolution space (11 of 14 were perfectly localized, while 3 of 14 were shifted by 1 voxel), and stage two demonstrated successful pituitary segmentation (anterior/posterior pituitary Dice_anterior_ = 0.97±0.02, Dice_posterior_ = 0.97±0.03, combined [total] gland Dice = 0.97±0.01). In an independent validation set (n = 40), segmentation overlap with adjudicated labels remained high (Dice_anterior_ = 0.91±0.04, Dice_posterior_ = 0.86±0.07, Dice_total_ = 0.93±0.03; Pearson correlation R_volume,anterior_=0.94, R_volume,posterior_=0.75, R_volume,combined_=0.95; Fig. 1). Further, in a linear model, there was no discernable systematic bias by visit, sex, or MRI scanner vendor (Dice_anterior_: p_visit_ = 0.40, p_sex_ = 0.14, p_vendor_ = 0.40). To contextualize this performance against established methods, we directly benchmarked the ABCD-specific network against FreeSurfer’s pituitary segmentation. The ABCD-specific neural net demonstrated higher overlap with adjudicated anterior-lobe labels in exter al validation (n=40; ABCD-specific anterior lobe Dice = 0.91±0.04; FreeSurfer anterior lobe Dice = 0.86±0.04, t_paired_=5.6, p<10^-5^), with manual inspection revealing occasional label spillover from FreeSurfer into the hypophyseal artery region. Despite this difference in spatial overlap, we note that FreeSurfer performance was comparable to that previously reported in adults (Anterior 0.83±0.04 (36)), anterior volumes were strongly correlated across methods in the full ABCD sample (n=30,276; R_anterior_=0.90), and primary associations were largely concordant across methods (see Supplementary Materials S1).

**Figure 1.**
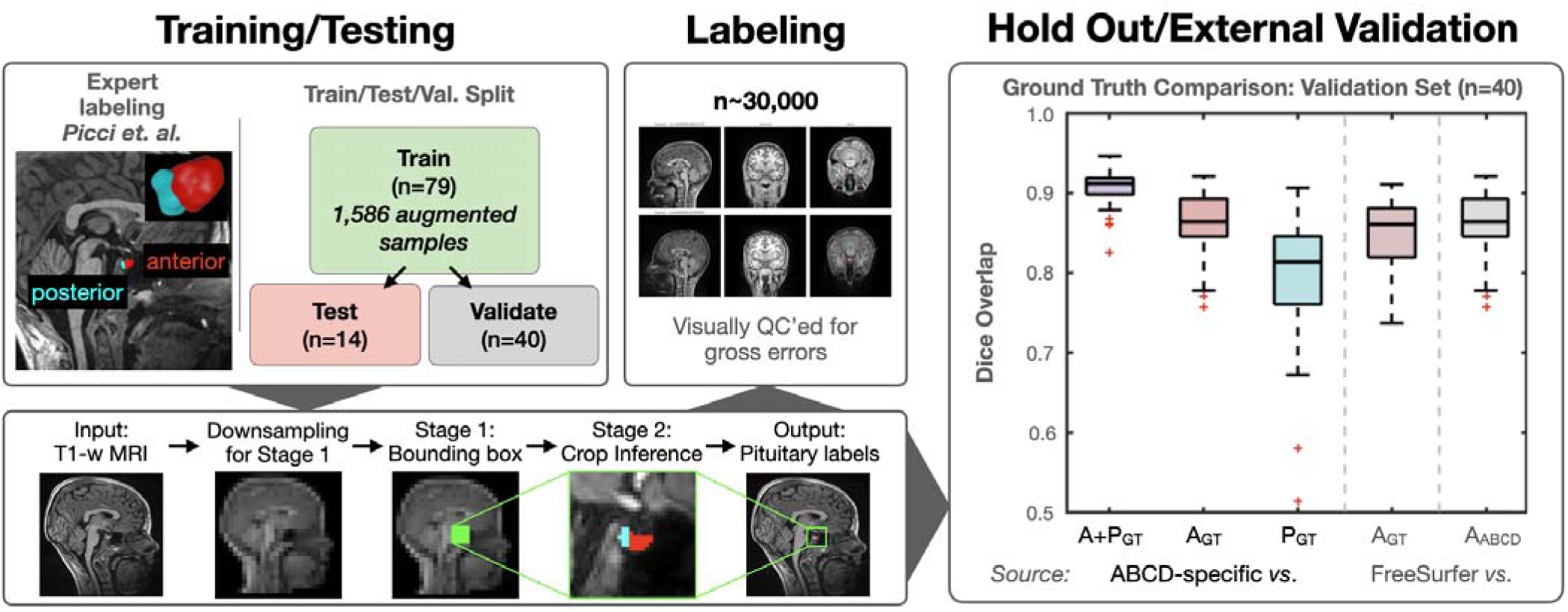
ABCD-specific neural network segmentation of the human anterior pituitary gland during adolescence. Left panels: A two-stage 3D U-Net was trained (n=79/14/40 train/test/validate split) to provide automated labeling of the anterior and posterior pituitary gland. Stage one downsamples and localizes a center-of-mass centered around the pituitary for cropping in preparation for stage two. Stage two provides anterior and posterior pituitary labels and projects the cropped box back into native space. Right panels: Dice performance is displayed for ABCD-specific segmentations against ground truth (GT) for combined (A+P), anterior (A), and posterior (P) pituitary. FreeSurfer comparisons demonstrate Dice scores in anterior pituitary against ground truth and ABCD-specific segmentations. Red crosses denote values outside of interquartile range.

To determine if the automated segmentations capture stable biological traits over time (up to 4 time points, across up to ∼6 years following baseline), we evaluated within-participant volume stability using generalized additive mixed models (ME-GAMMs) with random intercepts for participant, family, and site. After accounting for sex-specific age effects, substantial stable between-participant variability remained (participant SD = 44.7 mm³; residual SD = 49.9 mm³). This corresponds to a moderate conditional intraclass correlation (ICC = 0.44), suggesting that the extracted measures reflect stable individual traits and supporting the pipeline’s utility for large-scale developmental characterization.

### 2.2 Normative trajectories and sex differences in pituitary growth

In sex-stratified ME-GAMMs, anterior pituitary volume increased non-linearly with age in both males and females (males: estimated degrees of freedom (edf, a measure of “wiggliness”) = 7.26, F = 3,220, p < 10^-10^; females: edf = 7.79, F = 4,522, p < 10^-10^). Model comparison favored sex-specific over non-sex-specific (common) age curves (AIC_common_ = 342,656, AIC_sex-specific_ = 340,141, ΔAIC = 2,516 in favor of the sex-specific model), supporting distinct developmental trajectories (Fig. 2). A formal difference smooth confirmed that female and male age trajectories differed significantly in shape (edf = 7.11, F = 134.14, p < 10⁻¹⁰). Females showed earlier and steeper increases in anterior pituitary volume, with peak growth occurring at 11.45 years, whereas males showed a more protracted trajectory with peak growth at 13.03 years, and a maximal male–female difference in pituitary volume at 12.76 years. After adjusting for intracranial volume, the overall pattern of sex-differentiated growth trajectories was preserved, with the largest female–male difference at age 12.76 years (β = 102.7, SE = 2.49, t(df = 30,251) = 41.2, p < 10^-10^; Fig. 2). Across the observed age range, residual variability around the normative trajectories was greater in females than in males (residual SD: females = 87.0 mm³, males = 73.3 mm³, ratio = 1.19; F(df_num_ = 14,225, df_den_ = 16,049) = 1.41, p < 10^-10^; Fig. 3a). This pattern suggests a broader distribution of pituitary phenotypes among females, consistent with greater inter-individual variability in pubertal timing rendering chronological age a less precise predictor of pituitary volume in this group. Finally, supplementary analyses confirmed that ABCD-specific and FreeSurfer-based approaches produced highly consistent pituitary gland growth trajectories, while demonstrating that age-related variance is substantially more pronounced in the anterior compared to the posterior pituitary.

**Figure 2.**
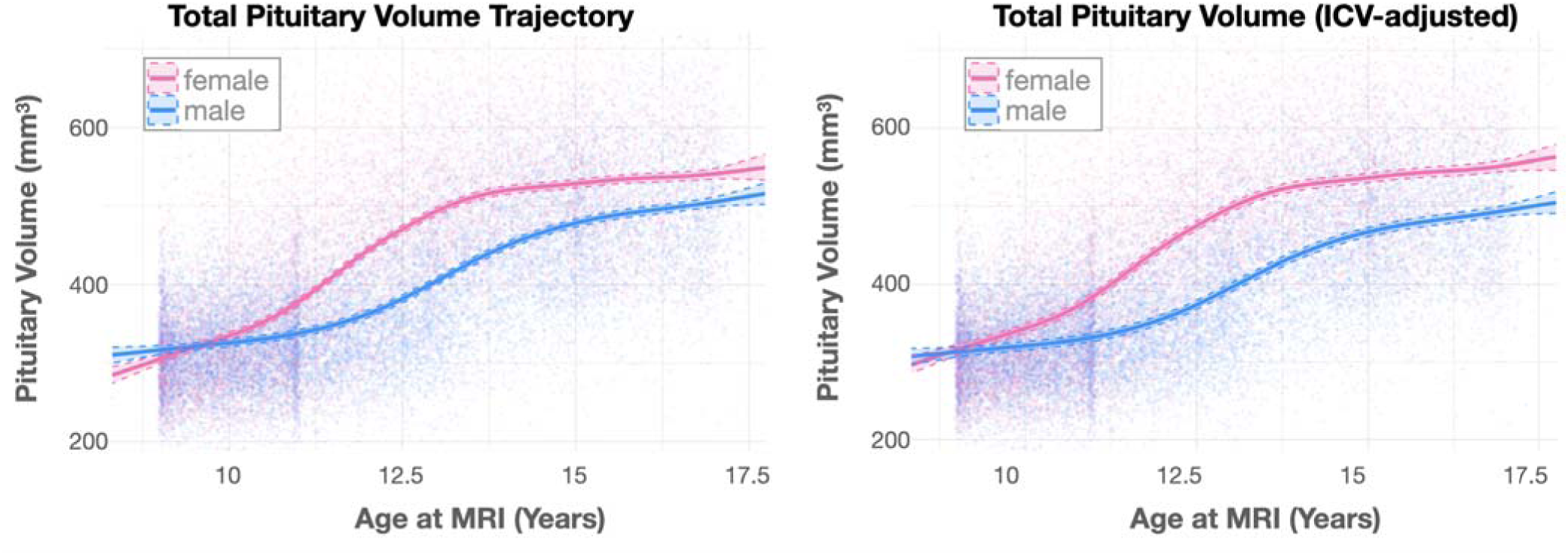
Developmental longitudinal trajectories of the human anterior pituitary gland during adolescence. Developmental trajectories, modeled using GAMMs, revealed sex-specific non-linear increases across development (left), including after adjustment for global size (intracranial volume, ICV).

**Figure 3.**
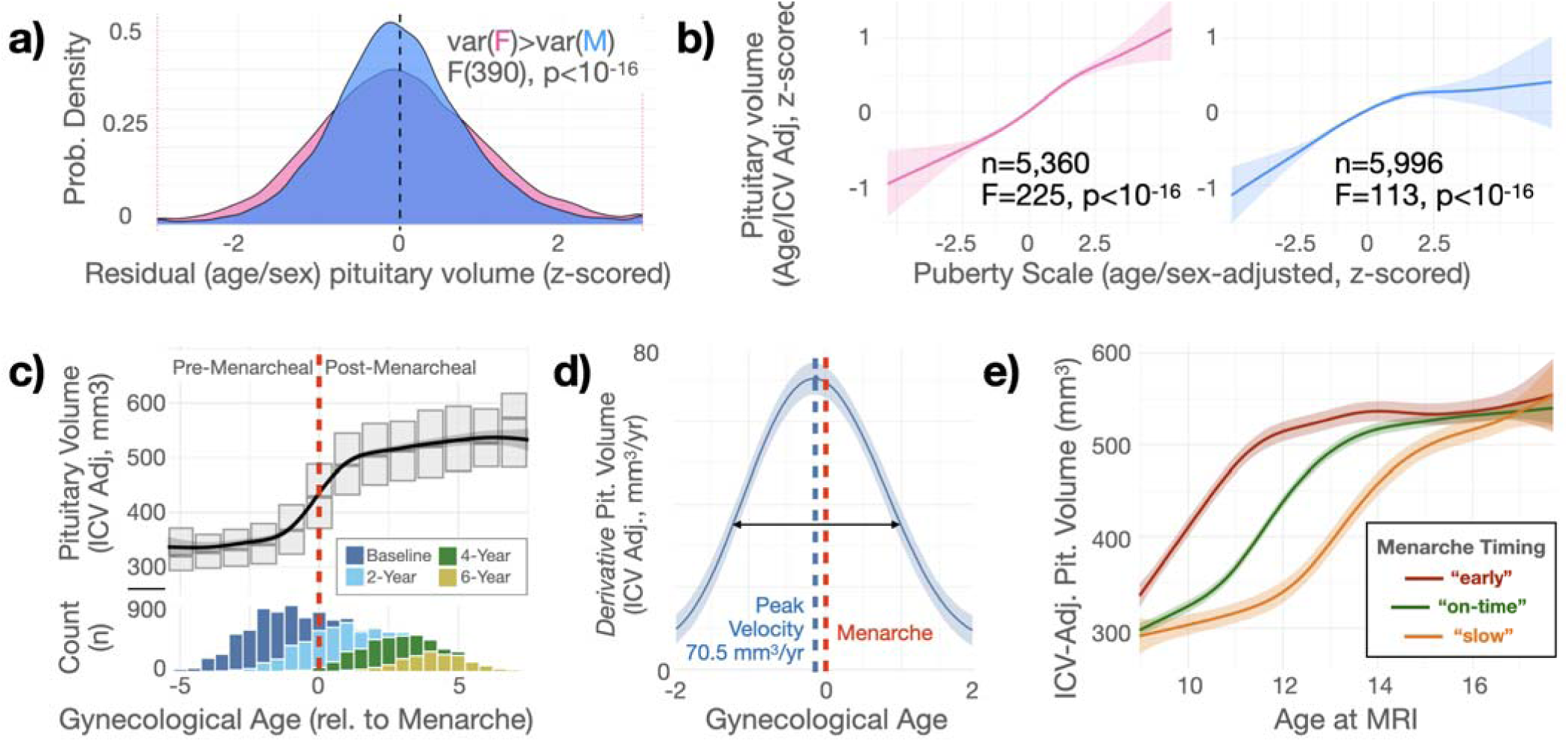
Anterior pituitary volume, pubertal tempo, and menarche onset. (a) Distribution of age- and sex-adjusted standardized residual anterior pituitary volume, showing greater variability in females than males. (b) Sex-stratified associations between pubertal maturation (puberty scale; age/sex-adjusted) and anterior pituitary volume (age/sex/ICV-adjusted), with shaded bands indicating model uncertainty. (c) Relationship between gynecological age at MRI and adjusted anterior pituitary volume, with age distribution by visit shown (counts). (d) Temporal derivative of the relationship between gynecological age at MRI and adjusted anterior pituitary volume identifies a maximal growth window centered on menarche (FWHM = 2.2 years, black arrow); the peak rate of change coincided with reported menarche age. (e) Longitudinal age-at-MRI trajectories of ICV-adjusted pituitary volume stratified by menarche timing group, illustrating larger and earlier increases in pituitary volume for “early” (<10.5yrs, n=1,363) menarche and delayed increases for “late” (>13yrs, n=991) menarche, relative to normal (*i.e*., 5^th^ to 95^th^ percentile) or “on-time” (n=10,865) menarche.

### 2.3 Anterior pituitary structure is associated with pubertal tempo and reproductive onset

We next examined whether volume of the anterior pituitary, the source of trophic hormones regulating gonadal hormone production, tracks pubertal development beyond age. After adjusting for age and intracranial volume, anterior pituitary volume increased with pubertal advancement (common smooth: edf = 6.44, F = 255, p < 10^-10^). Allowing the puberty association to differ by sex improved model fit relative to a common smooth (χ²(2) = 98.3, p < 10^-10^), and the relationship was positive in both sexes but stronger and more linear in females than males (edf_Female_=4.1, F_Female_=295; edf_Male_=5.4, F_Male_=104; Fig. 3b).

Youth-reported age at menarche was used as an objective developmental milestone to further investigate the link between anterior pituitary volume and reproductive maturation in females. First, across longitudinal visits, age- and ICV-adjusted anterior pituitary residuals were higher at post-menarcheal than pre-menarcheal assessments (Δ_mean_ = 0.36±0.02 S.E., t(df = 13,257) = 20.6, p <10^-10^, d=0.36). Second, using gynecological age (chronological age minus age at menarche) as a continuous frame of reference relative to menarche, we observed that anterior pituitary volume (adjusted for ICV) was non-linearly associated with gynecological age (edf = 8.75, F = 3400.3, p < 10⁻¹⁰; Fig. 3c). Further, a model using gynecological age and ICV only, was more parsimonious and explained more variance than a comparative model using chronological age, PDS, and ICV (ΔAIC = 631; R² = 0.558 vs. 0.527), suggesting that menarche-anchored developmental time (gynecological age) captures pituitary-relevant maturational variance that chronological age and pubertal staging together cannot fully resolve. Next, we used a temporal derivative of the fitted trajectory and posterior sampling to estimate a maximal growth window (FWHM=2.2 years, 95% CI: [2.1, 2.4], 125.5mm^3^ increase in volume roughly corresponding to 62.3% of total growth in adolescence, Fig. 3d). This window was centered on menarche: the peak rate of change coincided with reported menarche age (95% CI: [−0.73, +0.40] years, accounting for whole-year quantization in self-reported menarche; model-only CI [−0.19, −0.05] without this adjustment). Finally, to further quantify how different ages at menarche manifest in developmental trajectories, girls were grouped by menarche timing relative to the full sample (early, on-time, late); anterior pituitary trajectories differed across adolescence (Fig. 3e). A model allowing group-specific age smooths fit substantially better than a model with a shared age smooth (likelihood-ratio test: χ²(4) = 848.4, p < 10^-10^; ΔAIC = 857.0), further supporting pituitary morphology as a structural readout of the developing HPG axis. In the full model (adjusting for ICV and random effects of site, family, and participant), mean anterior pituitary volume also differed by timing group: early vs on-time β = 55.8 mm³ (SE = 3.3, t = 17.1, p < 10^-10^) and late vs on-time β = −45.6 mm³ (SE = 3.9, t = −11.7, p < 10^-10^). Supplementary analyses confirmed that all puberty and menarche associations were substantially stronger for the anterior than the posterior pituitary (Supplementary Materials S1). Together, these findings demonstrate that anterior pituitary volume is associated with pubertal maturation in both sexes, and in females, covaries with objective measures of menarche timing.

### 2.4. Shared and independent contributions of anterior pituitary and hormones

We next assessed the relative contributions of anterior pituitary volume and circulating sex steroid hormones to pubertal status and somatic growth, given that hormones are primary instigators driving changes in pubertal status (i.e., physical markers). Nested model comparisons confirmed that anterior pituitary volume provides unique information about pubertal status that is not captured by chronological age or circulating hormones alone (Table 1; Fig. 4). In females, the addition of anterior pituitary volume to a model containing age and hormones improved model fit (ΔAIC=−1,019). Permutation importance analysis identified anterior pituitary volume as the second most influential predictor of female pubertal status (after age), accounting for a reduction in explained variance (permutation importance = 0.22) when omitted relative to the contribution of any individual hormone (Fig. 4b). In males, while the full model including pituitary also outperformed the hormone-only model (ΔAIC=−380), the drivers of maturation were distinct from females. Specifically, while anterior pituitary volume remained a significant unique predictor, permutation importance indicated that male pubertal status was driven (after age) primarily by circulating testosterone (permutation importance = 0.20) followed by pituitary structure (permutation importance = 0.09; Fig. 4b).

**Figure 4.**
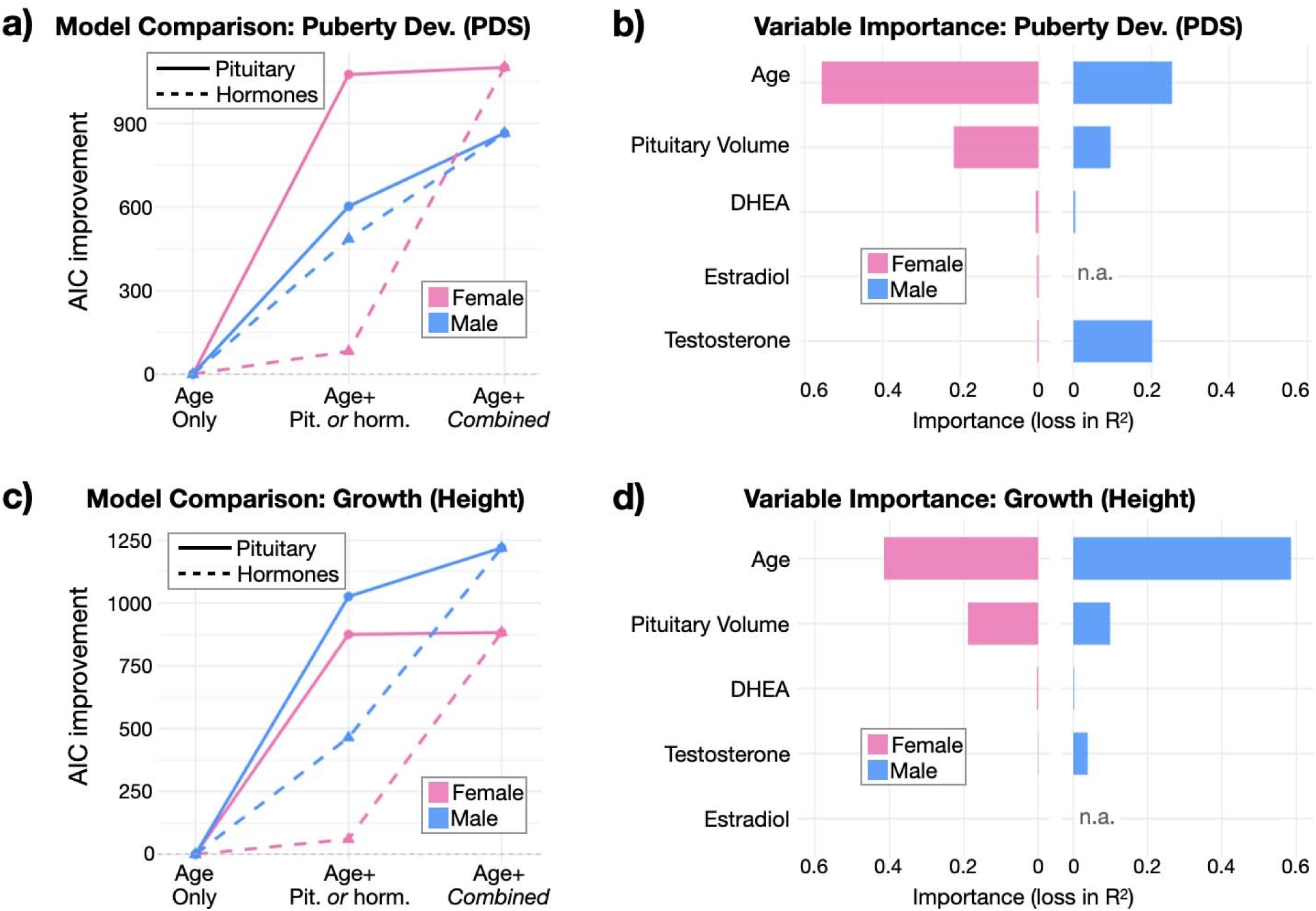
Distinct regulatory architectures in males and females. (a,c) Improvement in model fit (ΔAIC relative to an age-only reference model) when adding either anterior pituitary volume (solid lines) or hormones (dashed lines), or both (Combined), shown separately for females (pink) and males (blue) for (a) Pubertal Development Scale (PDS) and (c) height. (b,d) Relative premutation importance, quantified as average loss in explained variance (ΔR²) after randomly permuting each predictor within sex, for models modeling (c) PDS and (d) height. Age and anterior pituitary volume account for substantial unique variance in both outcomes, with sex-specific contributions of gonadal/adrenal hormones; estradiol terms are shown only for females.

**Table 1.** Distinct regulatory architectures in males and females: PDS. Generalized Additive Mixed Effects Models (GAMMs) linking pituitary volume and gonadal/adrenal hormones to pubertal development. Models shown summarize sex-stratified generalized additive mixed models of pubertal development (PDS) using age, pituitary volume, and circulating hormones, with smooth-term significance reported as estimated degrees of freedom (edf), F-statistics, and p-values. Model fit (adjusted R²) and sample sizes are provided for each sex; estradiol terms are included only in female models due to a lack of collection in males.

**Full model predicting PDS (Age + Pituitary + Hormones)**
| Predictor | Female edf | Female F | Female p | Male edf | Male F | Male p |
| --- | --- | --- | --- | --- | --- | --- |
| Age | 6.44 | 395.39 | < 1e-10 | 5.60 | 92.19 | < 1e-10 |
| Pituitary Volume | 6.30 | 183.44 | < 1e-10 | 5.93 | 70.71 | < 1e-10 |
| DHEA | 3.40 | 8.95 | 3.45e-06 | 3.63 | 6.68 | 6.64e-05 |
| Testosterone | 1.00 | 13.10 | 0.000297 | 6.56 | 41.99 | < 1e-10 |
| Estradiol | 2.32 | 9.13 | 5.69e-05 | — | — | — |

**Females:** Adj. $R^2$ = 0.69; N = 5,804 obs (3,809 subjects); **Males:** Adj. $R^2$ = 0.59; N = 7,089 obs (4,561 subjects)

This pattern of sex differences shifted when modeling somatic growth (Table 2). In contrast to PDS, residualized anterior pituitary volume was significantly associated with height (similarly residualized for age) in both females (permutation importance = 0.19) and males (permutation importance = 0.10). Notably, while testosterone remained a relevant predictor of height in males (permutation importance = 0.04), estradiol showed effectively zero association with height in females (permutation importance <0.001), demonstrating the model’s specificity in dissociating pubertal maturation from general somatic growth.

**Table 2.** Distinct regulatory architectures in males and females: Height. Generalized Additive Mixed Effects Models (GAMMs) linking pituitary volume and gonadal/adrenal hormones to height. Models shown summarize sex-stratified generalized additive mixed models of height using age, pituitary volume, and circulating hormones, with smooth-term significance reported as estimated degrees of freedom (edf), F-statistics, and p-values. Model fit (adjusted R²) and sample sizes are provided for each sex; estradiol terms are included only in female models due to a lack of collection in males.

**Full model predicting Height (Age + Pituitary + Hormones)**
| Predictor | Female edf | Female F | Female p | Male edf | Male F | Male p |
| --- | --- | --- | --- | --- | --- | --- |
| Age | 5.40 | 322.52 | < 1e-10 | 5.86 | 631.65 | < 1e-10 |
| Pituitary Volume | 5.50 | 168.12 | < 1e-10 | 5.19 | 158.57 | < 1e-10 |
| DHEA | 3.59 | 8.00 | 1.06e-05 | 4.47 | 6.10 | 4.70e-05 |
| Testosterone | 1.00 | 3.54 | 0.0601 | 6.99 | 28.77 | < 1e-10 |
| Estradiol | 1.00 | 0.16 | 0.6928 | — | — | — |

**Females:** Adj. $R^2 = 0.544$ ; N = 5,804 obs (3,809 subjects) **Males:** Adj. $R^2 = 0.728$ ; N = 7,089 obs (4,561 subjects)

### 2.5. Environmental and pharmacologic modulation of anterior pituitary structure

We next examined environmental stress and pharmacological influences on anterior pituitary volume. Adverse childhood experiences (ACEs) were associated with larger anterior pituitary volume after accounting for age, sex, and intracranial volume (Fig. 5a). In models treating ACEs as a continuous count variable (0–4+), higher ACE scores were associated with anterior pituitary residuals (β per ACE unit = 1.88, SE = 0.34, t(27,284) = 5.53, p < 10⁻⁷). When modeled categorically, youth with high ACE exposure (3+ events) showed larger anterior pituitary residuals relative to the low-ACE group (Δ_mean_ = 5.1mm^3^, SE = 0.99, t(df = 27,285) = 5.1, p < 10^-7^, d = 0.14).

**Figure 5.**
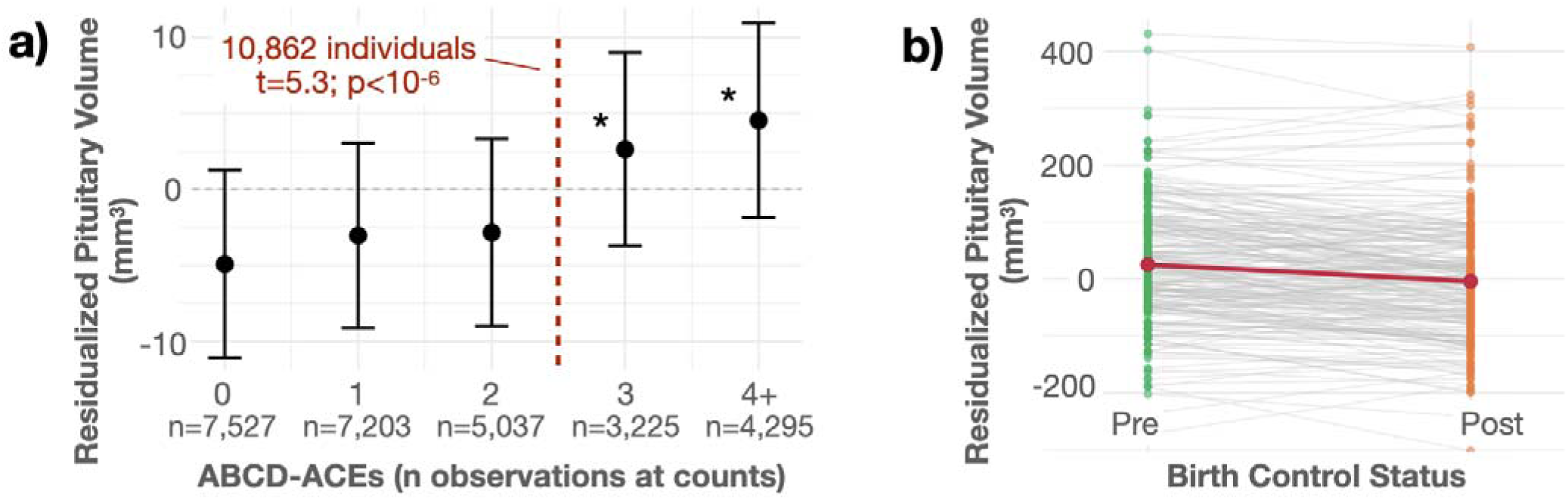
Adverse childhood experiences and anterior pituitary volume, and within-person change with birth control initiation. (a) Residualized anterior pituitary volume (age-, sex-, and ICV-adjusted) increases monotonically with cumulative ABCD adverse childhood experiences (ACEs; 0, 1, 2, 3, 4+), with group means and 95% CIs shown; sample sizes per ACE count are indicated below the x-axis. The dashed vertical line marks the literature-based high-exposure threshold (≥3 ACEs). Statistics shown reflect group low/high exposure comparisons. Error bars reflect 95% confidence intervals. (b) Within-person change in residualized anterior pituitary volume from pre- to post-initiation of hormonal birth control. Gray lines depict individual trajectories, points show observed values, and the overlaid red line indicates the estimated mean change across participants.

Because ACEs covary with socioeconomic disadvantage, we repeated these models adjusting for household income, caregiver education, and area deprivation index. The ACEs effect attenuated but remained significant (high-risk: Δ_mean_ = 3.78, SE = 1.07, t(23,648) = 3.53, p < 0.001), while each SES indicator was independently associated with pituitary volume (all p < 0.002) and directionally consistent with negative environments being associated with larger anterior pituitary volume. The full environmental model explained 1.8% of variance in pituitary residuals (a roughly six-fold increase over ACEs alone), supporting an effect broad early life stressors that is large relative to ACEs alone, but small relative to associations with puberty (ΔR² = 2.7–6.6%) and height (ΔR² = 3.1–4.8%).

Hormonal contraception was associated with cross-sectional and within-person differences in anterior pituitary residuals among females (Fig. 5b). In cross-sectional GAMMs, contraceptive use was related to smaller anterior pituitary residuals using both parent report (n = 4,184, β = -32.4, SE = 5.86, t = -5.53, p < 10^-7^) and youth report (n = 5,841, β = -24.5, SE = 3.18, t = -7.68, p < 10^-10^). Within-subject analyses identified n=238 females who transitioned from non-use to use over follow-up and contributed paired scans. Paired t-tests on nearest-neighbor pre–post scan pairs (age adjusted) yielded consistent results (t(df = 237) = -7.12, p < 10^-10^, d = -0.46). Together with the ACE findings, these models indicate that anterior pituitary structure is sensitive to both adverse environmental exposures and exogenous hormonal modulation in adolescence. Notably, neither ACEs nor hormonal contraception were significantly associated with posterior pituitary volume (Supplementary Materials S1), further supporting anatomical specificity of these environmental effects.

## 3. Discussion

Puberty is a major developmental transition during which neuroendocrine systems activate and reorganize with lasting consequences for growth, brain development, behavior, and health (37, 38), yet progress in understanding the neuroendocrine architecture underlying this transition has been limited by the lack of scalable in vivo measures of the pituitary, a central regulator of pubertal development. By leveraging deep learning at the scale of the ABCD Study, we demonstrate that the anterior pituitary undergoes systematic structural change across adolescence while preserving reliable individual differences. We mapped normative anterior pituitary growth across the pubertal transition and showed marked sex-specific differences in timing, tempo, and inter-individual variability. Anterior pituitary volume tracked pubertal maturation across both sexes; in females, pituitary volume changes were tightly linked to reproductive development, such that when indexed relative to menarche using gynecological age, pituitary development followed a non-linear trajectory with peak growth centered on menarche. Consistent with this, longitudinal pituitary growth trajectories differed among early-, on-time, and late-maturing groups, with earlier-maturing individuals showing earlier and larger increases in pituitary volume. Pituitary volume also captured variance in pubertal status and height beyond age and adjusted salivary hormones in females and males. Crucially, marked sex-specific differences emerged in the relative importance of hormones versus structure, thereby uncovering distinct regulatory architectures of pubertal maturation in females and males. Finally, anterior pituitary volume showed sensitivity to external modulation, increasing with cumulative adverse childhood experiences, and, decreasing cross-sectionally and within-person following initiation of hormonal contraception. Together, these findings suggest that anterior pituitary volume is a scalable, biologically grounded MRI-based marker associated with pubertal maturation and neuroendocrine plasticity during adolescence.

Anterior pituitary volume followed sex-specific, non-linear developmental trajectories, with peak pituitary growth occurring ∼1.5 years earlier in females compared to males, as well as more protracted expansion in males. Beyond mean differences, females showed substantial interindividual variability relative to males, underscoring the importance of large longitudinal samples for precisely estimating heterogeneity in pituitary-based indices of pubertal timing and tempo. To date, cross-sectional studies have consistently reported that females tend to have larger pituitary volume than males during pubertal development (15, 17, 19, 24, 39) and beyond (40). Longitudinal MRI studies of pituitary growth across adolescence remain limited. To our knowledge, only one prior study (n=249; 409 observations) explicitly modeled sex-specific normative developmental trajectories in a typically developing cohort (16). We extend this work by leveraging a population-scale, geographically diverse sample, enabling more precise estimation of sex differences in developmental timing, tempo, and interindividual variability. Further, pituitary development was tightly organized around reproductive development in females, such that when examined relative to menarche (i.e., gynecological age) (41), trajectories followed a non-linear pattern characterized by gradual increases that accelerated approaching menarche, with peak growth centered around menarche. Notably, the menarche-anchored reference frame captured more variance than models based on chronological age and pubertal status alone. Consistent with this, longitudinal trajectories differed across menarche timing groups (early, on-time, late), indicating that interindividual variation in menarcheal timing is reflected in distinct pituitary growth patterns. Note that these findings were robust to SES effects known to influence menarche timing as well as other indices of pubertal timing (42, 43). These findings extend prior work linking pituitary growth to pubertal maturation (16) by showing that pituitary morphology is associated with interindividual variation in pubertal timing and tempo longitudinally across adolescence. Larger anterior pituitary volumes in females may reflect earlier activation of hypothalamic-pituitary-axis signaling and estrogen-related trophic influences on pituitary tissue. Altogether, these findings position the anterior pituitary as a biologically proximal marker of HP axis timing and pubertal tempo.

We observed sex-differentiated predictive architectures of pubertal maturation: in males, circulating testosterone accounted for substantially more variance in pubertal status, whereas in females, anterior pituitary volume contributed more strongly, even after accounting for age and adjusted salivary hormones. Pituitary volume contributed independently in both sexes and improved model fit when added to age-only and hormone-only specifications, supporting nonredundant information carried by morphology and endocrine assays. Pituitary volume also predicted height in both sexes, whereas estradiol showed little association with height in females, underscoring dissociable contributions of structural and hormonal indices to distinct components of maturation. One plausible explanation is that estradiol levels are highly variable and cycle-dependent, even early in puberty prior to menarche (44), making single–time point measures a weaker index of developmental progression. In addition, concentrations of estradiol detected in salivary measures tend to be quite low (∼1-7% of total in circulation) (45). It may also be the case that estradiol alone is not a sufficient marker of somatic growth and pubertal maturation females, and that a broader panel of endocrine markers (e.g., LH, FSH, testosterone, progesterone) is necessary to more fully characterize pubertal development. In this context, pituitary structure may serve as a systems-level marker of endocrine capacity that is less sensitive to state-dependent fluctuations than estradiol assessed at any given visit, consistent with modest correspondence between estradiol levels and physical indices of pubertal maturation in early adolescent girls (46). In contrast, testosterone increases more monotonically across puberty in boys and shows stronger coupling with secondary sex characteristics (46, 47), such that circulating testosterone may track pubertal status more directly in males, with pituitary structure providing complementary but less dominant information. Taken together, these patterns suggest that anterior pituitary volume functions as a global, biologically specific MRI-derived index that complements subjective pubertal staging and peripheral endocrine measures, consistent with prior ABCD work demonstrating nonredundant contributions of hormones and pubertal staging (48). More broadly, structural and hormonal markers may index pubertal maturation on partially distinct timescales, with pituitary morphology reflecting more integrated neuroendocrine change and circulating hormones capturing more state-dependent signaling.

Greater exposure to adverse childhood experiences was associated with larger anterior pituitary volume in adolescence, evident both as an approximately linear increase with each additional exposure and as higher volumes among youth with the highest cumulative adversity (e.g., ≥3 types). This pattern extends prior reports from smaller samples indicating that pituitary morphology is sensitive to adverse contexts during development (22, 23, 25, 49). Importantly, these adversity-related elevations were observed against a backdrop of normative pubertal growth in the broader cohort, suggesting that environmental stressors may shift pituitary structure beyond what would be expected from (appreciably larger) age- and puberty-related changes alone. Such shifts are broadly aligned with stress acceleration frameworks (50, 51), which posit that early adversity may recalibrate neuroendocrine maturation toward earlier or heightened engagement under conditions of threat or unpredictability, providing population-scale neuroimaging evidence for neuroendocrine mechanisms proposed by these models. Thus, anterior pituitary structure may provide a neurobiological index linking early environmental adversity to altered pubertal trajectories that have long been documented in developmental literature (35). One interpretation is that the anterior pituitary, as a central hub linking hypothalamic control to multiple peripheral endocrine axes, reflects cumulative regulatory load across adolescence. Chronic stress exposure is known to increase trophic signaling through the HPA axis, leading to sustained stimulation of corticotroph populations that may contribute to volumetric expansion, consistent with prior literature examining effects of early life stress on pituitary volume (25, 26). Such sustained activation may also engage intra-pituitary signaling such that local cross-talk, including paracrine growth-factor signaling, propagates these effects beyond a single cell type (e.g., corticotrophs) to other cell populations, consistent with broadly distributed volumetric shifts rather than a narrowly axis-specific change. The adversity construct examined here captures a broad range of exposures including maltreatment, material deprivation, family separation, and neighborhood safety (52), supporting the view that pituitary morphology may index aggregate neuroendocrine burden associated with early environmental stress. Further, the association between adversity exposure and anterior pituitary volume remained significant after adjusting for multiple SES factors, demonstrating that the observed adversity effects were not explained by SES disadvantage alone. Notably, greater socioeconomic disadvantage was also independently associated with larger anterior pituitary volume (R² increasing from 0.3% to 1.8%), and the adversity-specific association persisted after SES adjustment. That two distinct but stress-linked dimensions of the early environment (cumulative adversity and socioeconomic disadvantage) converge on the same structural signature reinforces the interpretation that the anterior pituitary is sensitive to stress as a broad construct, rather than to any single exposure type.

Hormonal contraceptive use was associated with smaller anterior pituitary volume, evident both cross-sectionally and as within-person reductions following initiation, indicating that pituitary structure remains responsive to pharmacologic modulation during adolescence. Although specific formulations were not available here, most combined estrogen-progestin contraceptives suppress endogenous HPG-axis activity by reducing gonadotropin-releasing hormone-dependent secretion of luteinizing hormone and follicle-stimulating hormone, thereby dampening endogenous ovarian hormone production (53). Reduced HPG-axis drive may lower trophic demand on anterior pituitary cell populations over time, providing a plausible mechanism for the volumetric decreases observed. Importantly, the within-person change following initiation strengthens interpretation by reducing confounding from stable individual differences and aligns with prior *adult* studies reporting smaller pituitary volume among oral contraceptive users (54–56). Together, these findings support the view that the anterior pituitary shows structural changes associated with shifting regulatory demands during adolescence rather than being a fixed anatomical structure.

Anterior pituitary volume provides an objective, biologically grounded MRI-derived marker of pubertal maturation that complements, rather than simply recapitulates, circulating hormone assays. Rather than serving as a surrogate for any single hormone, pituitary morphology likely reflects integrated neuroendocrine remodeling within the anterior lobe, with tissue-level change unfolding on longer developmental timescales than state-dependent salivary hormone snapshots. Even under careful preprocessing, salivary hormones remain sensitive to collection and handling factors (*e.g.,* timing relative to waking, clock time, recent meal, caffeine, exercise, and storage conditions), motivating extensive de-nuisancing to reduce measurement noise while preserving developmental signal. In this context, pituitary volume is biologically proximal to hypothalamic regulatory signals and may index cumulative endocrine demand across adolescence through coordinated changes in anterior pituitary cell populations (*e.g.,* gonadotropes, somatotropes, lactotropes). As a result, structural measures of the anterior pituitary may ultimately provide a useful imaging marker for identifying atypical neuroendocrine development or risk trajectories for puberty- or menstrual-related health conditions. This image-based phenotype therefore enables new tests of brain–body coupling in ABCD, including how endocrine maturation relates to trajectories of brain development, physical health, and behavior. Pituitary-derived measures may also be a powerful complementary measure for estimating pubertal development in neuroimaging datasets where hormone assays or staging assessments are unavailable. In keeping with the goals of population neuroscience, derived measures can be readily recreated from publicly shared code (*see Data Availability*) and shared through established ABCD data-access pathways.

The current work featured a developmentally sensitive ABCD-specific neural network for hypothesis testing. While this approach has clear advantages, the recently introduced FreeSurfer pituitary module conceptually represents a tradeoff between cohort-specific accuracy and broad portability. Specifically, it provides an accessible, standardized alternative, but like many automated pipelines it is anchored in adult anatomy and may not be expected to capture the rapid morphological change and local signal variation that characterize adolescence. Conversely, bespoke approaches like the one used here can achieve high performance in-distribution yet fail silently when confronted with unseen acquisition characteristics or anatomy. This motivates a reciprocal validation strategy: study-specific models can be benchmarked against a widely used external method, while simultaneously providing evidence about whether a general-purpose tool is sufficiently accurate in pediatric samples. In our external validation, the ABCD-specific model showed higher overlap with adjudicated anterior-lobe labels (Dice 0.91 vs 0.86). However, volumes were strongly correlated with one another across the full ABCD sample, and, most importantly, primary associations were largely concordant across methods. Collectively, on the basis of evidence, we suggest that either approach is suitable for use in the ABCD study. Further, we believe that there may be promise in their combined use. For example, method disagreement can flag images/labels for targeted image censoring, and ensembling may further stabilize estimates for hypothesis testing in the ABCD study.

Despite many strengths of this study, several limitations should be considered when interpreting these findings. Hormonal measures were repeated across waves but derived from single time-point salivary assays at each visit. Thus, even when de-nuisanced, single time-point assays are subject to substantial state dependence and measurement error, which likely attenuates our estimates of associations between hormones and pituitary development. Replication in cohorts with denser endocrine sampling will be important to reduce attenuation from state dependence and measurement error and to yield more precise effect-size estimates of hormone–pituitary coupling. Moreover, hidden confounding cannot be ruled out, including overlapping genetic influences and unmeasured axis-level factors (*e.g.,* growth axis and its hormones, hypothalamic or gonadal development (57)) that may jointly shape pituitary morphology and pubertal or somatic maturation. Pituitary volume is a coarse, macroscopic marker that does not necessarily reflect processes at the cellular level driving endocrine function, limiting axis- and cell-type specificity. Multimodal MRI, including quantitative approaches, may improve sensitivity to microstructural or compositional change beyond what is captured by gross volume, but mechanistic interpretation will likely require integration with experimental and cross-species work that can directly assay endocrine cell populations and signaling dynamics. In parallel, environmental effects should be refined beyond simple cumulative adversity counts to test whether specific dimensions of stress (*e.g.,* timing, chronicity, caregiver buffering, unpredictability) differentially shape neuroendocrine development, consistent with emerging cross-species evidence linking unpredictability to stress-sensitive circuitry (58, 59).

This work describes the anterior pituitary as a scalable, biologically grounded neuroendocrine transducer that integrates pubertal maturation and environmental experience into a stable structural phenotype relevant to behavioral development. By leveraging deep learning on a population scale, we provide a framework moving beyond the limitations of subjective staging and volatile hormone measurements to map precise, sex-specific trajectories of endocrine tempo. The observed sexual dimorphism (characterized by earlier, more variable, menarche-anchored, and structurally dominant pituitary growth in females) reveals distinct regulatory architectures for pubertal maturation that are not captured by circulating hormones alone. Furthermore, the sensitivity of pituitary volume to both cumulative adversity and pharmacologic modulation underscores its plasticity as a target of biological embedding during adolescence, a period of heightened behavioral vulnerability and change. Collectively, these findings further position the pituitary as a dynamic, tractable biomarker for large-scale neuroimaging, bridging the gap between central neuroendocrine signaling, behavioral development, and environmental health.

## 4. Material and Methods

### 4.1 Sample Description

This work used data from the ABCD Study release 6.0 (https://doi.org/10.82525/jy7n-g441). ABCD is a multisite longitudinal study of youth followed from late childhood into adolescence (∼9-17 years old) designed to capture a representative sample of sociodemographic variation (see Compton et. al. for a more complete discussion of representativeness) from 2017 to present. We considered structural MRI and phenotypic data from this release, using minimally preprocessed T1-weighted images and associated variables accessed through the LASSO query platform and downloaded using Globus. Participants were eligible for inclusion in this analysis if they had at least one usable (suggested minimally preprocessed T1s) T1-weighted scan and complete data for age, sex at birth, site, family structure, and intracranial volume. Derived pituitary volumes (see below for details) were available for all participants in the analytic sample. This final sample comprised 11,818 individuals contributing 30,276 observations across four visits. At baseline, 11,755 participants contributed data (10.0 ± 0.6 years, 6,139 male and 5,616 female), with 8,092 at the 2-year follow-up (12.0 ± 0.7 years, 4,362 male and 3,730 female), 6,343 at the 4-year follow-up (14.2 ± 0.7 years, 3,403 male and 2,940 female), and 4,086 at the 6-year follow-up (16.1 ± 0.7 years, 2,146 male and 1,940 female). The apparent reduction in participants at later visits is expected to primarily reflect the staggered release of data rather than attrition; many participants have not yet reached these visits due to differences in enrollment timing (early estimates of study withdrawal are low, ∼0.01%) (60). Ethical review for ABCD study data collection has been described elsewhere, and a non-human subjects self-determination was filed with the local University of California Irvine Institutional Review Board for the current secondary analysis.

### 4.2 Training and Test Data: Manual Labeling and Data Augmentation

Manual pituitary segmentations were generated by an expert rater (GP) using a previously validated protocol (GP) (15, 22, 61). Specifically, these tracing procedures have previously demonstrated high interrater reliability in volume (i.e., intraclass correlation coefficients ∼0.93–0.97) (15). In the present study, the anterior and posterior lobes of the pituitary gland were traced on n=94 baseline T1-weighted images using 3D Slicer [v. 5.6.2, (62)]. Labels were constructed in coronal and sagittal planes, using established anatomical landmarks. Superior boundaries were defined by the diaphragm sellae and third ventricle, the sphenoid sinus and bony sella inferiorly, and the internal carotid arteries bilaterally. The anterior and posterior lobes were distinguished in the sagittal plane based on T1-weighted posterior hyperintensity.

Data augmentation with rigid-registration was used to increase the effective sample size for training while preserving pituitary morphology. First, manual labels were split into training (n=80) and test (n=14) partitions. This split was chosen to maximize the sample available for training purposes, while having a plausible sample size for testing purposes (see below for further validation details). For augmentation, the 80 training images were treated as “moving” images and rigidly aligned to 20 randomly chosen “target” images (per training image, drawn from the same 80) using ANTs’ *antsRegistrationSyNQuick.sh*. This design yielded up to 1,600 possible moving–target combinations with ABCD-specific contrast and anatomy. Of these, 1,586 registrations completed successfully and were retained for model training; one moving image consistently failed to register and was excluded, resulting in 1,586 augmented image–label pairs derived from 79 original labeled scans. Label interpolation was performed using ANTs’ *GenericLabel[Linear]* parameter. Notably, because only rigid transforms were applied to both images and labels, augmentation introduced variation in global head position and orientation and modest variation in label borders/edges (due to interpolation), while preserving the local shape and anatomy of the pituitary relative to surrounding structures.

### 4.3 Two-stage pipeline overview: pituitary localization and lobar segmentation

We implemented a two-stage pipeline to first localize (stage 1) and then segment the pituitary in a reduced field of view (stage 2). This design reduces GPU memory demands to fit on commercially available hardware, mitigates extreme class imbalance between pituitary tissue and background, and forces the model to focus on structures surrounding the pituitary. In the first stage, a coarse localization model was trained to predict a cubic region of interest (32 × 32 × 32 mm) surrounding the geometric pituitary center of mass in a space downsampled to an 8 mm isotropic resolution. The stage 1 model was trained to predict this binary box in this downsampled space.

In the second stage, a high-resolution (1mm isotropic native) lobar segmentation model was trained to segment the anterior and posterior pituitary within the small field of view. To match the distribution of inputs at test time, these cropped training images were centered using the pituitary location predicted by the trained stage 1 model rather than the manual center of mass. The stage 2 model takes the native-resolution 32 × 32 × 32 crop as input and produces a three-class segmentation (background, anterior lobe, posterior lobe) that can then be mapped back into the full field-of-view. Collectively, at inference, this two-stage model then firsts crops the T1w image, segments the anterior/posterior pituitary, and remaps the small field-of-view back into native space (<3s total time on a NVIDIA 1080 GPU for inference).

### 4.4 U-Net architecture, training parameters, and hardware

Each of the two stages were trained using a 3D U-Net architecture. Networks were implemented in PyTorch (v2.5.0+cu124) with supporting libraries NumPy (v2.1.2) and NiBabel (v5.3.2). Each model consisted of three down-sampling and three up-sampling levels with skip connections between corresponding encoder and decoder layers, Instance Normalization, and ReLU activations. The stage 1 model produced a single-channel output corresponding to the pituitary localization box, whereas the stage 2 model produced a three-channel output corresponding to background, anterior lobe, and posterior lobe. Models were trained on a single NVIDIA GTX 1080 GPU, with training for both stages taking approximately 36 hours in total.

Models were trained for 1,000 epochs, batch size of 2, and using an Adam optimizer with a fixed learning rate of 1×10⁻⁴. This epoch count was chosen because validation performance plateaued well before 1,000 epochs (with good test performance evident by ∼100 epochs), and there was no evidence of overfitting, as reflected by continued decreases in training loss without degradation in test validation metrics. For the localization (stage 1) model, we used a Dice loss to optimize prediction of the pituitary box. For the lobar segmentation (stage 2) model, we used a multi-class cross-entropy loss with three labels (0 = background, 1 = anterior lobe, 2 = posterior lobe). Hyperparameters were selected based on commonly used values in 3D U-Net applications rather than an extensive hyperparameter search. The resulting models are tuned to the characteristics of ABCD T1-weighted data; while preliminary tests on other datasets suggest some degree of generalizability, we do not guarantee or assume full portability to other cohorts or imaging protocols.

### 4.5 Network Validation

Network performance was evaluated in three ways. First, we defined an independent validation set of 40 T1-weighted images balanced by visit (10 each at the baseline, 2-year, 4-year, and 6-year follow-ups), sex (20 male, 20 female), and site (at least one T1-weighted image from each site), randomly selected to meet these criteria. This validation set was manually segmented using the protocol described above and compared with the two-stage network output using Sørensen–Dice coefficients of overlap. Second, all T1-weighted images included in the study were visualized in three orthogonal views with the T1-weighted image as underlay and derived labels as overlay, combined into a single PDF document. This document was reviewed for gross errors and provided as Supplementary Material. Lastly, we provide downstream scientific validation by testing a set of a priori hypotheses that depend on accurate pituitary segmentation, including developmental trajectories and associations with hormones, pubertal status, and environmental measures previously associated with pituitary gland volume and growth (14–16, 18, 23). Together, these complementary validations aim to support the robustness of the automated pituitary segmentation across the ABCD sample.

### 4.6 Anthropometric and pubertal measures

Height and weight were obtained from ABCD release 6.0 (https://doi.org/10.82525/jy7n-g441) tabulated data (*ph_y_anthr__height_mean*, *ph_y_anthr__weight_mean*). Youth report pubertal status was indexed using ABCD’s composite Pubertal Development Scale (PDS) scores. Sex-specific PDS composites were used for females (*ph_y_pds_f_mean*) and males (*ph_y_pds_m_mean*). Because these composites are constructed on different criteria, nearly all puberty related analyses were conducted on a stratified basis. Limited exceptions testing for sex-specific effects and trajectories (i.e., interactions) used within-sex z-scored equivalents and a unified continuous PDS created.

In females, menarche status and age at menarche were derived from both caregiver- and youth-reported items. Binary indicators of menarche (*ph_p_pds__f_002*, *ph_y_pds__f_002*) were recoded as “Yes” versus “No,” with non-informative responses (e.g., “n/a,” “don’t know”) treated as missing. Reported age at menarche was based on both parent and youth report (*ph_p_pds__f_002__01*, *ph_y_pds__f_002__01*). For participants with multiple reports, we defined a participant-level age at menarche as the median of reported measures. Event-time measures relative to menarche were then computed as scan age minus participant-level menarche age and used in models examining pituitary trajectories before and after menarche (see Statistical Analyses). Further, girls were categorized as early (<10.5), on-time (10.5-13), or late (>13) based on the observed distribution of menarche age.

### 4.7 Salivary hormone measures

Salivary dehydroepiandrosterone (DHEA), testosterone, and estradiol were obtained from longitudinal hormone assays included in ABCD release 6.0 (*ph_y_phs__dhea_mean, ph_y_phs__ert_mean, ph_y_phs__hse_mean*). Mean hormone concentrations were transformed using base-2 logarithms to reduce skew and bring values towards a more normal distribution. Hormones were *a priori* de-nuisanced (see below) using a set of saliva collection and handling variables: age at saliva collection (*ph_y_phs_age*), clock time and timing relative to waking (*ph_y_phs_001*, *ph_y_phs__coll_t*, *ph_y_phs__coll__end_t*), time since last meal (*ph_y_phs__lastmeal_hr*), month of collection (*ph_y_phs_dtt*, treated cyclically), freezer temperature (*ph_y_phs__freeze_001*), time to freezer (ph_y_phs__freeze_t), recent caffeine intake (*ph_y_phs_003*), recent exercise (*ph_y_phs_004*), and concern about sample quality (*ph_y_phs_001*). Non-numeric entries (e.g., “n/a”) were set to missing, and time-of-day variables were converted to continuous minute-based measures where applicable.

To derive hormone measures that preserve age-related developmental signal while reducing measurement noise, we implemented a two-step “de-nuisancing” procedure. First, for each hormone we screened individual collection variables using univariate generalized additive models (GAMs) to identify relevant non-linear and categorical predictors. Second, for each hormone we fit a multivariable generalized additive mixed model (GAMM) in participants with at least two observations, modeling log2 hormone levels as a function of: (i) smooth terms for continuous collection-related predictors, (ii) a cyclic smooth term for month of collection, (iii) categorical terms for freezer temperature, sample concern, caffeine intake, and exercise, and (iv) random intercepts for participant, family, and site. Age at saliva collection was modeled as a distinct smooth term whose contribution was explicitly preserved. From each fitted model we obtained residuals from the mixed-effects component and the model-based age-related smooth. “Adjusted” hormone values were then constructed as the sum of the grand mean, residual, and age smooth, yielding age-preserved, de-nuisanced log2 hormone measures. This procedure was applied in the full sample and repeated stratified by sex. Sex-stratified, age-preserved adjusted hormone variables served as the primary hormone measures in downstream analyses.

### 4.8 External Exposures: Adverse childhood experiences and hormonal contraception

Adverse childhood experiences (ACEs) were operationalized using an ABCD-specific ACEs proxy score (ABCD-ACEs) based on prior work in the ABCD cohort (52). Following that framework to extend measures from Year 2 to Year 6, we identified adversity-related items spanning core ACE domains and implemented in-house scoring in R using publicly available code (52). Items were harmonized across reporters (youth, caregiver) and combined into a count of endorsed adversities with truncated score where values ≥4 were collapsed into a “4+” category to roughly balance groups for analysis. Several adjustments were made in computing our ACEs proxy score to accommodate differences between the 5.1 and 6.0 ABCD data releases. First, the Kiddie Schedule for Affective Disorders (KSADS) was shared as a total sum in 6.0, as opposed to the raw variable scores, which were required for addition of specific questions to certain domains of our ACES proxy score, therefore we used KSADS data from the 5.1 release. Additionally, KSADS scores from Year 2 were repeated for later ACEs calculations because this data was not provided in following years. Adult Self-Report (parent mental illness and substance abuse) score was given as Total Problems ASR Syndrome Scale (t-score) T-scores in 5.1, but this was no longer available in the 6.0 release. We instead utilized a threshold <64 on the ASR scale to generate the binary (yes/no) score for parental mental illness, supported by previous work with this scale (CITE https://pmc.ncbi.nlm.nih.gov/articles/PMC7441087/). Family history of mental illness was originally calculated including parental history, but the new data release included sibling mental illness history which was added to the updated ACEs proxy score. Lastly, the Child’s Report of Parental Behavior Inventory (CRPBI) was updated to include new survey data at all years of collected, except for the emotional neglect item with was not available at Year 2.

Socioeconomic status was indexed using three indicators: household income, highest caregiver education, and area deprivation index (ADI). Baseline ADI values were carried forward across sessions within participant. Each indicator was z-scored, with ADI inverted so that higher values consistently indicated higher SES. A composite SES score was computed as the mean of the three z-scored indicators; a median split on this composite defined High and Low SES groups for descriptive and moderation analyses. In ACEs sensitivity models, the three raw indicators (income, education, ADI) were entered as separate parametric covariates. In puberty and menarche sensitivity analyses, the composite score was used as a single linear covariate.

Hormonal contraception exposure was derived from both youth-reported and caregiver-reported items indicating current use of hormonal birth control (ph_y_pds__f_002__05, ph_p_pds__f_002__05). For cross-sectional analyses, youth- and parent-reported contraception were modeled separately to assess consistency across informants. For within-subject analyses, we constructed a combined contraception-status variable that classified participants as using hormonal contraception if either reporter endorsed current use (“Yes”), and as not using if no reports indicated use and at least one report indicated non-use (“No”). Among females with longitudinal data, we then identified participants who transitioned from non-use to use over time and defined a within-person period variable contrasting observations before versus after the first reported onset of hormonal contraception. This combined and transition-based coding was used to test within-subject changes in pituitary volume associated with initiating hormonal contraception.

### 4.9 Statistical Analyses

Longitudinal statistical analyses were conducted to understand normative trajectories of pituitary volume across adolescence, puberty- and growth-specific associations with pituitary volume, and the role of external influences on pituitary volume. All pituitary volume outcomes were winsorized at ±3 SD prior to analysis to limit the influence of extreme values; both raw and winsorized values were retained, with winsorized values used in all reported models.

To characterize normative trajectories of pituitary growth, we fit generalized additive mixed models (ME-GAMMs) predicting anterior pituitary volume from age and sex (all smooths specified as thin-plate regression splines, default k=10). Sex was modeled as a fixed effect, and age-related growth was modeled using sex-specific smooth terms, specifying a GAMM of the form *∼ sex + s(age, by = sex)*, with random intercepts for site, family, and participant to account for clustering and repeated measures. This model was repeated using non-sex-specific smooths to compare fits via Akaike Information Criterion (AIC) metrics. To directly test the age-by-sex interaction, we additionally fit a GAM with an ordered factor coding for sex and a difference smooth *(∼ sex + s(age) + s(age, by = sex_ordered)*). Peak growth velocity was determined using numerical differentiation. We then fit an ICV-adjusted normative model that additionally included sex-specific smooth terms for intracranial volume, *volume ∼ sex + s(age, by = sex) + s(icv, by = sex)*, with the same random-effects structure. Sex-differences in adjusted and unadjusted models were compared at the age of maximum pituitary volume difference in the unadjusted model in order to bias differences towards the unadjusted model (and thereby removing any bias towards the adjusted model). All models were estimated using the gamm4 package in R v4.5.2. Model-based predictions and 95% confidence intervals were generated over the observed age range separately for males and females; for the ICV-adjusted model, trajectories were evaluated at the sample mean ICV to visualize sex-specific normative growth curves at a common head-size reference. Concurvity diagnostics for all GAMMs indicated low to moderate non-linear dependence among predictors (worst-case concurvity indices for age- and ICV-related smooth terms < 0.45).

Our primary puberty models estimated the association between relative pubertal status for age and relative pituitary volume for age separately in males and females. Specifically, we fit a GAMM of the form volume_resid_ ∼ s(pds_resid_) + s(ICV_resid_) + random intercepts for site, family structure, and participant. Here (and throughout) volume_resid_ is pituitary volume residualized for age and is used as the outcome rather than the predictor for more normally distributed error values. This parameterization allows non-linear associations between pubertal advancement and pituitary volume, while adjusting for individual differences in head size beyond age. Model-based predictions and 95% confidence intervals were generated over the observed range of PDS in each sex, holding ICV_resid_ at their sex-specific means, and were visualized separately for males and females.

To complement PDS scores, we leveraged youth-reported age at menarche as a discrete, biologically anchored marker of reproductive onset. First, age- and ICV-adjusted pituitary residuals were compared between post-menarcheal and pre-menarcheal visits using a GAMM with a fixed effect of menarche status and random intercepts for site, family, and participant. To characterize pituitary trajectories relative to menarche, we computed gynecological age as scan age minus participant-level menarche age for all female observations with a known menarche age, such that negative values indicate pre-menarcheal scans and positive values indicate post-menarcheal scans. Using all available longitudinal data (N = 13,219 observations from 5,012 individuals across four sessions), we fit a GAMM with raw anterior pituitary volume as the outcome, smooth terms for gynecological age and ICV (each with thin-plate regression spline bases), and random intercepts for site, family, and participant. In addition, we evaluated the first derivative of the gynecological age smooth to identify the point of maximum volumetric change, with 95% confidence intervals on the peak location obtained via posterior simulation (1,000 draws from the fitted coefficient distribution). The full width at half maximum (FWHM) of the derivative curve and the proportion of total predicted growth within it were estimated on the same draws. Because self-reported menarche age is recorded in whole years, we also report a quantization-adjusted CI for the peak (SD = 1/√12 yr on the reference point) to conservatively account for measurement error. To assess whether gynecological age provides a more informative developmental axis than conventional pubertal staging, we compared this model against a model using chronological age, PDS, and ICV on an identical subsample requiring non-missing PDS (N = 11,891 observations from 4,844 individuals), using AIC and marginal R². Finally, we defined categorical groups of menarche timing roughly representative of the bottom and top 10 percentile values, and the middle 80 percentile as follows: early (<10.5 years), on-time (10.5–13 years), and late (>13 years). Using all available longitudinal data from females, we fit a GAMM with pituitary volume as the outcome, fixed effects for menarche tempo group, separate age smooths by group, a smooth term for ICV, and random intercepts for site, family, and participant [pituitary ∼ menarche_group + s(age, by = menarche_group) + s(icv)]. Predicted trajectories and 95% confidence intervals were generated on a grid spanning the observed age range with ICV held at its sample mean, and visualized together with the raw longitudinal data to illustrate pituitary growth profiles for Fast, On-time, and Slow menarche groups.

To understand the unique contribution of variance in pituitary volume to pubertal scores, over and above hormone concentration measurements, we constructed a set of nested GAMMs. A parallel set of GAMMs were identically constructed using height as an outcome, due to its objective measurement relative to the PDS scale. Note that we used within sample normative modeling here, as opposed to CDC measures used in supplementary materials, in order to make measures and models as comparable between height and PDS as possible. As detailed above, salivary hormone measures were first de-nuisanced using GAMM-based adjustment to preserve age-related signal while removing collection-related noise. Anterior pituitary volume was reduced to a single z-scored residual index by regressing volume on sex-specific smooth terms for age and ICV in a GAMM (with random intercepts for site, family, and participant) and z-scoring the residuals from the mixed-effects (lmer) component to facilitate comparability between PDS and height models. Similarly, PDS, height, and hormones were z-scored nested by sex and hormone. For each sex and outcome (PDS and height), we then fit a set of nested GAMMs with random intercepts for site, family, and participant. First, using PDS/height as outcomes, we fit smooths to: pituitary only, hormones only, and pituitary and hormones jointly. Additional female hormone models were ran dropping estradiol from the model for comparability with males lacking estradiol assays.

To determine whether pituitary volume provides information about pubertal status over and above circulating hormone concentrations, we constructed a series of nested Generalized Additive Mixed Models (GAMMs) with random intercepts for site, family, and participant. Analyses were performed for two outcomes: Pubertal Development Scale (PDS) and height, treating the latter as an objective anthropometric validator of somatic growth. Prior to modeling, salivary hormone measures (DHEA, testosterone, and estradiol) were adjusted for nuisance factors using a GAMM-based approach to preserve age-related signal while removing collection-related noise (see above). For each sex and outcome, we fit four nested models to evaluate the unique and joint contributions of pituitary structure and hormonal function: a baseline model containing age only; models adding either pituitary volume or hormones separately; and a full model containing age, pituitary volume, and hormones. All continuous predictors were modeled using penalized thin-plate regression splines to accommodate non-linear developmental trajectories. We evaluated the specific contribution of pituitary volume using two complementary approaches. First, we compared nested models using the Akaike Information Criterion (AIC) and Likelihood Ratio Tests (LRT), defining unique contribution as a significant improvement in model fit (lower AIC, p<.05) when adding pituitary volume to a model already containing age and hormones. Second, to quantify the predictive power of specific features within the full model, we calculated permutation importance (63), which measures the reduction in explained variance (R²) when a single predictor’s values are randomly shuffled, thereby breaking its association with the outcome while preserving the structure of all other predictors. This metric then isolates the unique contribution of each predictor.

Finally, to understand the role of external exposures, GAMMs were constructed using pituitary volume as an outcome and each exposure (ACEs, birth control) as predictors, in separate models. ACEs and birth control measures used are defined above. Four total models were tested. First, ACEs were examined both as an ordinal count (0–4+, with values ≥4 collapsed) and as a binary risk (0–2 vs 3+ events), using GAMMs consistent with the above. To assess whether ACEs effects were independent of socioeconomic context, ACEs models were repeated with household income, caregiver education, and area deprivation index as additional parametric covariates (N = 23,653 observations from 9,731 individuals with complete SES data). Next, hormonal contraception analyses were restricted to females and used contraceptive use as a fixed effect in an analogous GAMM. To quantify within-person change at contraceptive initiation, we used paired-scan linear mixed-effects models with random intercepts for participant. Each of these exposures are described more fully above. Multiple comparisons adjustment was done using Bonferroni throughout, within families of analyses.

### 4.10 Data Sharing

Underlying individual participant data and corresponding data dictionaries are shared as part of the ABCD data repository at the NIH Brain Development Cohorts (NBDC) Data Sharing Platform (https://www.nbdc-datahub.org/abcd-study). All variables used in this manuscript are denoted by their data dictionary name. Note that future ABCD data release variable names may not be consistent with those used here. All derived data (e.g., residualized pituitary volumes, cleaned hormone measures) can be recreated through public access code (https://github.com/jerodras/abcd_pituitary), and will be made available upon request. As with all ABCD data, necessary Data Use Agreements will be required for data access.

## Data Availability

All derived data (e.g., residualized pituitary volumes, cleaned hormone measures) can be recreated through public access code (https://github.com/jerodras/abcd_pituitary), and will be made available upon request. As with all ABCD data, necessary Data Use Agreements will be required for data access.

## Acknowledgements

We thank the participant volunteers for donating their time to the dHCP and ABCD projects and the generosity of the European Research Council and the National Institute of Health for making the dHCP and ABCD data publicly available.

Data used in the preparation of this article were obtained from the Adolescent Brain Cognitive Development^SM^ (ABCD) Study (abcdstudy.org), held in the NIMH Data Archive (NDA). This is a multisite, longitudinal study designed to recruit more than 10,000 children age 9-10 and follow them over 10 years into early adulthood. The ABCD Study® is supported by the National Institutes of Health and additional federal partners under award numbers U01DA041048, U01DA050989, U01DA051016, U01DA041022, U01DA051018, U01DA051037, U01DA050987, U01DA041174, U01DA041106, U01DA041117, U01DA041028, U01DA041134, U01DA050988, U01DA051039, U01DA041156, U01DA041025, U01DA041120, U01DA051038, U01DA041148, U01DA041093, U01DA041089, U24DA041123, U24DA041147. A full list of supporters is available at https://abcdstudy.org/federal-partners.html. A listing of participating sites and a complete listing of the study investigators can be found at https://abcdstudy.org/consortium_members/. ABCD consortium investigators designed and implemented the study and/or provided data but did not necessarily participate in the analysis or writing of this report. This manuscript reflects the views of the authors and may not reflect the opinions or views of the NIH or ABCD consortium investigators.

The ABCD data repository grows and changes over time. The ABCD data used in this report came from [NIMH Data Archive Digital Object Identifier (DOI): <u>10.15154/z563-zd24</u>]. DOIs can be found at [https://nda.nih.gov/abcd/abcd-annual-releases].

## Authors’ contributions

G.P. and J.M.R.: Conceptualization, Methodology, Formal analysis, Investigation, Writing, Visualization, Funding acquisition. D.M.B.: Resources, Writing, Supervision. B.L. and C.H.: Methodology, Formal analysis, Writing. D.A.F.: Conceptualization, Resources, Writing. B.T.L.: Data curation, Formal analysis. L.H.S.: Resources, Formal analysis, Writing. M.A.Y.: Resources, Writing. H.S. and L.R.W.: Formal analysis, Writing. E.A.S.: Conceptualization, Methodology, Writing.

## Prior Presentation

Portions of this work have been presented as a poster at the Flux Congress 2025 in Dublin, Ireland.

