## Supplementary Material for "Longitudinal MRI reveals adolescent pituitary growth patterns linked to puberty and environmental exposures"

*Supplementary S1.1: Multiverse Analyses Using Alternate Measures of Pituitary Volume (FreeSurfer and Custom ABCD-Specific Neural Network)*

In the main manuscript we use anterior pituitary volume measured using a custom ABCD-specific neural network for segmentation. The rationale for anterior pituitary is supported by its biological relevance as the primary gland for the synthesis and release of stimulating hormones that regulate adolescent puberty and growth. Here, as supplementary material, we repeat and report on primary analyses using a total of six different pituitary measures, including the anterior pituitary measure used in the main manuscript. The five additional measures are comprised of the posterior and combined/total gland volume from the custom neural net, and anterior, posterior, combined gland volume derived using the recently released pituitary neural network trained on adults and available as part of FreeSurfer software release 8.1. The subsections below are broken down to mirror the results sections in the main manuscript, specifically: Normative trajectories and sex differences in pituitary growth (Manuscript Section 2.2), pubertal tempo and reproductive onset (Manuscript Section 2.3), shared and independent contributions of pituitary and hormones (Manuscript Section 2.4), environmental and pharmacologic modulation of pituitary structure (Manuscript Section 2.5).

*Supplementary S1.2: Normative trajectories and sex differences in pituitary growth*

In the main manuscript we report sex-specific non-linear trajectories across adolescence. Here we repeat these analyses and provide basic statistics including full model fits (R^2^ for ICV-adjusted and unadjusted models), variance partitioning (conditional ICC for participant, family structure, and site), and non-linear sex-specific effects of age (edf and F-statistic) using the six different measures of pituitary volume to demonstrate the biological relevance of the anterior pituitary, and measurement validity through generalizable effects to external tools (FreeSurfer). While ABCD-specific pituitary segmentations had a slight advantage in total variance explained, reduced site-related variance, increased sex-differences in residual variance, and some larger effect sizes (especially posterior), the FreeSurfer segmentations had a slight edge in within-person stability across visits. However, both methods produced highly consistent findings in size, direction, and significance supporting the measurement validity of both approaches. With respect to anterior vs. posterior portions, anterior pituitary effects tended to be larger than posterior, and combined gland models were comparable to anterior models.

| **Pituitary Measure** | **R^2^_unadj_** | **R^2^_adj_** | **F_age,M_** | **F_age,F_** | **cICC_ind_** | **cICC_site_** | **cICC_fam_** | **varfm** |
| --- | --- | --- | --- | --- | --- | --- | --- | --- |
| **Anterior (Manuscript)** | **0.47** | **0.48** | 4424 | 3144 | 0.44 | 0.04 | 0.44 | **1.47** |
| **Posterior** | 0.17 | 0.19 | 1355 | 883 | **0.54** | **0.16** | 0.35 | 1.10 |
| **Combined** | 0.46 | 0.47 | 4556 | **3179** | 0.48 | 0.05 | 0.45 | 1.43 |
| **Anterior *FreeSurfer*** | 0.46 | 0.46 | 4499 | 2968 | 0.48 | 0.05 | 0.47 | 1.41 |
| **Posterior *FreeSurfer*** | 0.11 | 0.11 | 845 | 655 | **0.54** | 0.15 | 0.39 | 0.93 |
| **Combined *FreeSurfer*** | 0.45 | 0.45 | **4717** | 3148 | 0.51 | 0.08 | **0.48** | 1.41 |

*Supplementary S1.3: Pubertal tempo and reproductive onset*

In the main manuscript we report age-adjusted associations between anterior pituitary volume and puberty scores (PDS), and the model comparison effect size when allowing for sex-specific associations. Here we repeat these analyses and provide basic statistics including full model fits (R^2^), effect sizes for PDS (edf and F), and improvements in fit when using sex-specific curves (χ^2^) using the six different measures of pituitary volume. ABCD-specific pituitary segmentations had a slight advantage in total variance explained and effect size of association with PDS, FreeSurfer measures appeared more sensitive to sex-specific effects. However, both methods produced highly consistent findings in size, direction, and significance supporting the measurement validity of both approaches. With respect to anterior vs. posterior portions, anterior pituitary effects tended to be larger than posterior, and combined gland models were comparable to anterior models.

| **Pituitary Measure** | **R^2^_adj_** | **edf_pds_** | **F_pds_** | **χ^2^** |
| --- | --- | --- | --- | --- |
| **Anterior (Manuscript)** | **0.079** | 6.35 | **251.5** | 95.9 |
| **Posterior** | 0.014 | 4.05 | 64.4 | 9.9 |
| **Combined** | 0.076 | **6.88** | 221.2 | 83.4 |
| **Anterior FreeSurfer** | 0.073 | 6.50 | 235.7 | 134.9 |
| **Posterior FreeSurfer** | 0.017 | 1.00 | 193.2 | 29.0 |
| **Combined FreeSurfer** | 0.078 | 6.90 | 228.9 | **142.9** |

In the main manuscript we report associations between age/ICV-adjusted anterior pituitary volume at baseline and age at menarche, and model comparisons between pituitary growth trajectories classified by early/on-time/late menarche. Here we repeat these analyses and provide basic statistics including effect sizes for pituitary volume and binary classification of having reached menarche (t_yes/no_), effect sizes for baseline pituitary volume and age at menarche (edf and F), and improvements in fit when using menarche timing-specific curves (χ^2^) using the six different measures of pituitary volume. FreeSurfer measures demonstrated slightly stronger associations with age at menarche effects. However, both methods produced highly consistent findings in size, direction, and significance supporting the measurement validity of both approaches. With respect to anterior vs. posterior portions, anterior pituitary effects tended to be larger than posterior, and combined gland models were comparable to anterior models.

| **Pituitary Measure** | **t_yes/no_** | **edf_menarche,age_** | **F_menarche,age_** | **χ^2^** |
| --- | --- | --- | --- | --- |
| **Anterior (Manuscript)** | 20.3 | **6.19** | 52.9 | 809.4 |
| **Posterior** | 10.6 | 5.30 | 13.9 | 178.3 |
| **Combined** | 20.3 | 6.05 | 53.0 | 819.4 |
| **Anterior FreeSurfer** | **21.7** | 5.97 | 64.8 | 1036.5 |
| **Posterior FreeSurfer** | 6.9 | 3.42 | 16.2 | 181.0 |
| **Combined FreeSurfer** | 21.7 | 5.89 | **66.8** | **1107.9** |

*Supplementary S1.4: Shared and independent contributions of pituitary and hormones*

In the main manuscript we report associations between anterior pituitary volume and puberty scores (PDS) and somatic growth (height), while adjusting for salivary hormone concentrations to demonstrate the relevance of pituitary measures over and above (measured) circulating hormones. Here we repeat these analyses and report effect sizes for PDS and height (F_pds_ and F_height_) while adjusting for hormone concentrations using the six different measures of pituitary volume. As in the main manuscript, this is done stratified by sex based on the premise that these are sex-specific trajectories and regulatory systems, and the simple practical fact that estradiol measures are not included for males but expected to be important in female development. Thus, resulting in four separate sets of analyses (sex x outcome). Broadly, ABCD-specific pituitary segmentations demonstrated slightly higher effect sizes of association with PDS and height. However, both methods produced highly consistent findings in size, direction, and significance supporting the measurement validity of both approaches. With respect to anterior vs. posterior portions, anterior pituitary effects tended to be larger than posterior, and combined gland models were comparable to anterior models.

|  | **F_PDS_** | | **F_Height_** | |
| --- | --- | --- | --- | --- |
|  | **Male** | **Female** | **Male** | **Female** |
| **Anterior (Manuscript)** | **68.65** | **175.42** | **147.13** | **158.35** |
| **Posterior** | 32.95 | 24.36 | 34.45 | 29.9 |
| **Combined** | 58.21 | 165.59 | 128.47 | 133.33 |
| **Anterior FreeSurfer** | 56.38 | 149.9 | 122.15 | 133.31 |
| **Posterior FreeSurfer** | 50.1 | 34.41 | 40.87 | 37.13 |
| **Combined FreeSurfer** | 60.52 | 174.77 | 129.55 | 133.6 |

*Supplementary S1.5: Environmental and pharmacologic modulation of pituitary structure*

In the main manuscript we report associations between early life stress (ACEs) and anterior pituitary volume, and separately, the effect of oral contraception on anterior pituitary volume using paired (pre/post) observations. Here we repeat these analyses and report effect sizes for ACEs (t_ACEs_) while adjusting for sex-specific effects of age and ICV. In addition, in females, we report the pre/post effect size (t_oc_) while repeating analyses using the six different measures of pituitary volume. Contrary to the majority of multiverse analyses above, FreeSurfer demonstrated a small advantage in effect sizes of stress (ACEs) and oral contraceptive. However, also as above, both methods produced highly consistent findings in size, direction, and significance supporting the measurement validity of both approaches. With respect to anterior vs. posterior portions, anterior pituitary effects tended to be larger than posterior, and combined gland models were comparable to anterior models.

|  | **t_aces_** | **t_oc_** |
| --- | --- | --- |
| **Anterior (Manuscript)** | 5.07 | -6.93 |
| **Posterior** | 2.64 | -1.59 |
| **Combined** | 5.03 | -6.73 |
| **Anterior FreeSurfer** | **5.37** | -9.43 |
| **Posterior FreeSurfer** | 0.69 | -4.37 |
| **Combined FreeSurfer** | 5.05 | **-9.90** |

*Supplementary S2.1: Validation of Sex-Specific Age Trajectories in an Independent Cohort (HCP-D)*

This supplemental external validation analysis included longitudinal data from 1,269 youth (1,654 total observations) aged 5-22 years from the Human Connectome Project in Development (HCP-D). Participants were recruited from four sites: Harvard University, University of California-Los Angeles, University of Minnesota, and Washington University in St. Louis. The study was approved by the Institutional Review Board at Washington University in St. Louis. Participants provided written informed consent or assent, and parents of participants under 18 years old provided written informed consent for the child’s participation.

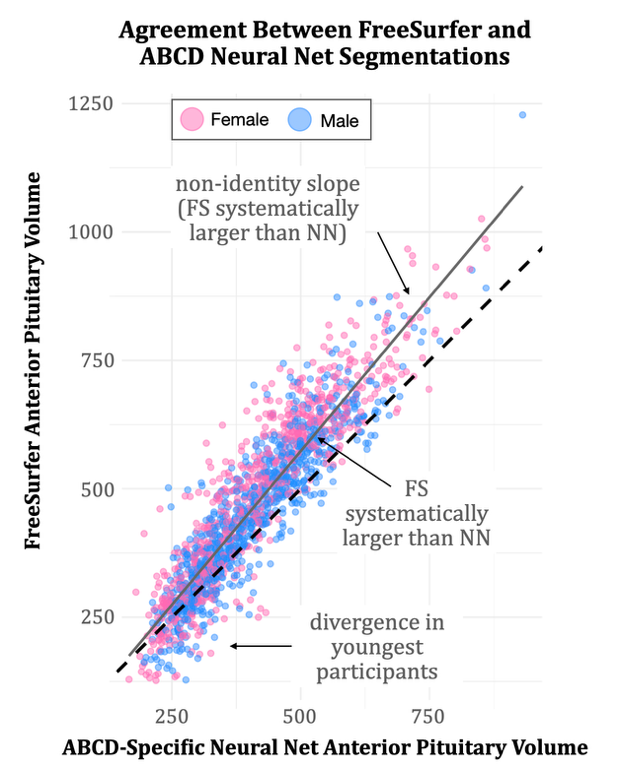
HCP-D T1-weighted images were downloaded from the ConnectomeDB Internal-Facing Site (IntraDB) using xnat_file_downloader.sh shell script to pull the most recent specific session MRI data available. Minimally pre-processed images were used to minimize site bias, and downsampled from the native 0.8mm isotropic resolution used in the HCP-D study to the 1.0mm space and orientation in ABCD to allow for inference using the ABCD-specific neural networks. Notably, because these networks were trained on ages, resolution, and contrasts that subset the HCP-D study, there is likely to be additional error in domain transfer. Thus, the effects reported here are expected to be conservative estimates in comparison to retraining on 0.8mm and HCP-D label image pairs. Otherwise, anterior and posterior volumes were derived identically to those reported in the main manuscript.

Automated ABCD-trained neural network (NN) segmentations of the anterior pituitary showed strong concordance with standard FreeSurfer (FS) estimates in the independent HCP-D cohort (Figure 1). Extreme disagreements between the two methods (>3 SD difference in residual volume) accounted for approximately 2% of the overall images and underwent strict manual visual inspection. This inspection identified systematic failure modes including incomplete or missing tissue segmentation and tissue hallucination, prompting the exclusion of these specific cases from further analysis. However, we note that at this failure rate, the inclusion of these outlying segmentations in sensitivity analyses (unreported) did not materially impact the below findings.

**Figure 1.** Agreement between FreeSurfer (FS) and ABCD-specific Neural Networks (NN).

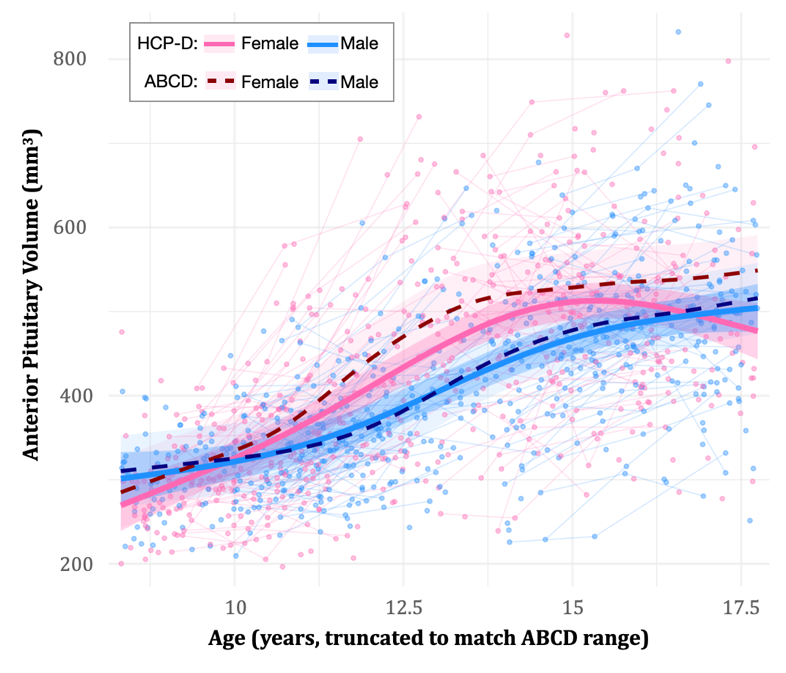
To test the generalizability of the developmental patterns observed in the reference ABCD cohort, sex-specific age trajectories (unadjusted for ICV) were derived in the HCP-D data using identical modeling procedures used for the ABCD cohort in the main manuscript. The finding of non-linear, sex-specific age trajectories in anterior pituitary, with girls preceding boys in growth, was successfully replicated in HCP-D (females: edf = 6.69, F = 139.66, p < 0.001; males: edf = 6.15, F = 160.88, p < 0.001). Furthermore, the mean developmental trajectory of the HCP-D cohort fell largely within the ±1 SD normative bounds of the ABCD reference model (90% overlap for females, 100% for males; Figure 2) within the overlapping age range. To formally test for cohort-specific trajectories, the two datasets were merged without prior harmonization and stratified by sex. Model comparison via AIC contrasted a shared-trajectory model against a model permitting a cohort-by-age interaction smooth. In both males and females, the shared-trajectory model was preferred (females: ΔAIC = +10.9 favoring null; males: ΔAIC = +4.3 favoring null), indicating no improvement from allowing cohort-specific age trajectories. The cohort-specific smooths in the alternative model were both highly non-linear (edf ≈ 6.5 per cohort), confirming that this result reflects genuine trajectory similarity rather than over-penalization. In males, there was no significant main effect of cohort on baseline volume (β = −4.98, SE = 5.70, t = −0.87, p = 0.382), whereas there was a minor decrease in HCP-D females (intercept ~5% smaller in HCP-D females; β = −22.38, SE = 8.96, t = −2.50, p = 0.015).

**Figure 2.** Sex-specific trajectories in the HCP-D Study are consistent with, and overlap those, in the ABCD Study.

*Supplementary S2.2: Generalizability of Growth-Related Phenotypes in an Independent Cohort (HCP-D)*

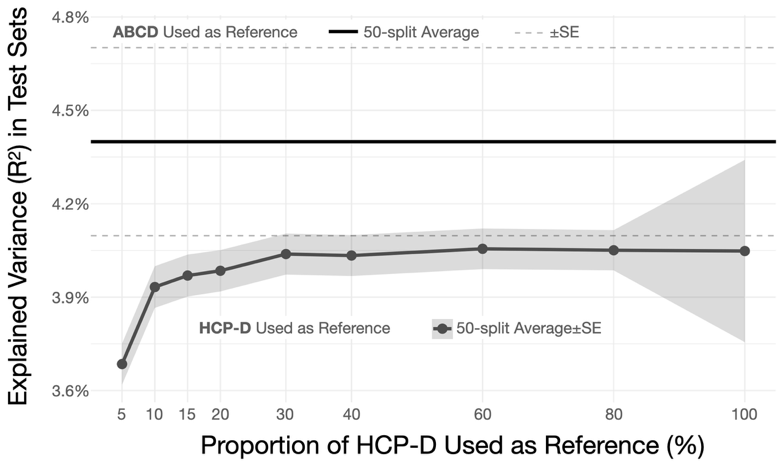
To assess the clinical and biological utility of the normative models, we evaluated the relationship between anterior pituitary volume and physical growth (standing height) in the HCP-D replication sample. Physical growth was standardized using CDC height-for-age reference curves (Z-scores) to ensure independence from both the ABCD and HCP-D neuroimaging models. We compared two distinct methods for deriving normalized pituitary volumes (Z-scores): 1) deriving residuals internally using age- and sex-specific generalized additive models (GAMs) fit directly to the local HCP-D sample, and 2) projecting the HCP-D subjects onto the pre-trained normative models derived from the massive ABCD reference cohort. We hypothesized that the ABCD reference model, benefiting from a vastly larger training sample, would yield more precise, generalizable estimates of expected volume, thereby producing residuals more strongly associated with concurrent deviations in height. We confirmed that while both measures were significantly associated with CDC height Z-scores (ABCD-derived residuals: r = 0.208, t(1166) = 7.26, p < 0.001; locally-derived residuals: r = 0.198–0.199, t(1166) ≈ 6.91, p < 0.001), the ABCD-derived residuals were significantly more so (Steiger's z = 2.32, p = 0.020 vs. local wiggly; z = 2.06, p = 0.040 vs. local regularized).

Finally, to explicitly demonstrate the mechanism driving this difference and to evaluate the effect of local sample size, we performed a robust Monte Carlo subsampling simulation. The HCP-D cohort was subjected to 50-fold repeated random sub-sampling validation. In each of the 50 iterations, a 20% holdout test set was isolated and completely withheld from model training. Local GAMs were then trained on incrementally increasing proportions of the remaining data pool (ranging from 5% to 100%) and subsequently forced to predict the expected volumes for the unseen subjects in the holdout test set. We found that locally fitted models exhibited substantial drops in out-of-sample explained variance (R²), a hallmark of the overfitting-to-underfitting tradeoff inherent to smaller sample sizes. Crucially, regardless of the proportion of local training data utilized, the local HCP-D models never achieved the out-of-sample biological validity (variance explained in height) provided by the fixed ABCD reference norms (Figure 3).

**Figure 3.** Using ABCD as a reference dataset improves associations between anterior pituitary volume and height-for-age relative to HCP-D as a self-reference.

Collectively, the above demonstrates that projecting data from smaller or moderately sized cohorts onto population-level normative models isolates more biologically meaningful variance than fitting models directly to the local sample. Conversely, if the primary objective is simply to establish a group-level statistical effect to inform developmental theory, fitting models to a smaller local sample remains perfectly adequate. This is supported by the observation that even in the smallest subset of training data (5%, n=48 in-sample) a significant association (p<0.05) was observed in the hold-out test set in 82% of the Monte Carlo samples. However, these results also highlight the clear advantages of adopting large-scale normative frameworks for clinical or precision-medicine applications, and the ability of the current neural networks to do so, where the reliability of individual-level phenotyping is paramount.

*Supplementary S3.1: Descriptive Statistics from the ABCD Dataset*

The ABCD Study imaging sample used in this study comprised 11,762 individuals at baseline, with participants returning at the 2-year, 4-year, and 6-year follow-up sessions, respectively (Supplementary Fig. 4). Age distributions shifted rightward across sessions as expected, spanning approximately 9–10 years at baseline to 15–18 years at the 6-year follow-up.

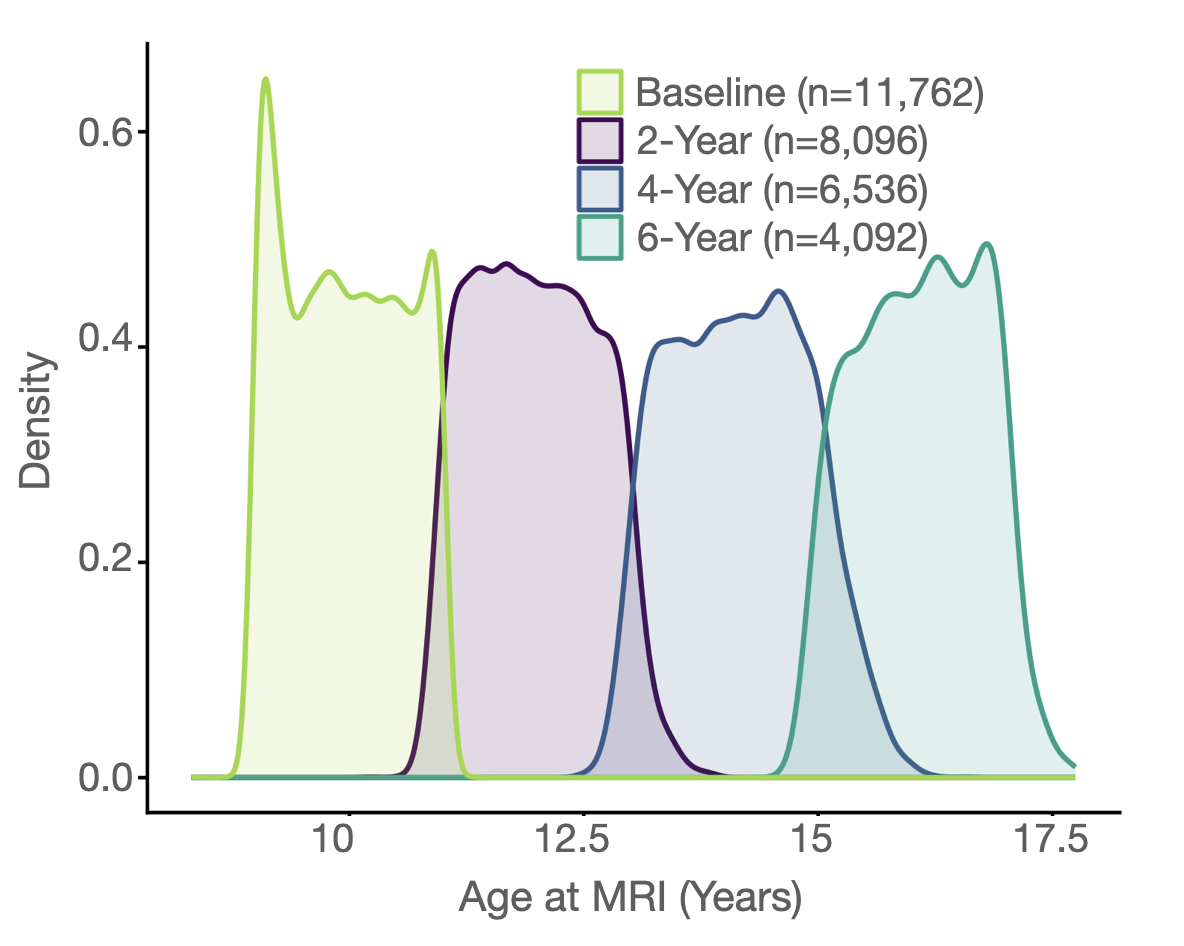

**Figure 4.** **Density plots broken down by visit.**

To characterize the developmental trajectories of salivary hormones available in the ABCD Study, sex-stratified GAMMs were fit for DHEA, testosterone, and estradiol (females only) as functions of age, with random intercepts for site, family, and participant (Supplementary Fig. 5). All hormones showed strongly nonlinear age trajectories in both sexes (all p < 10⁻¹⁰). DHEA increased steadily across adolescence in both sexes (female: edf = 6.9, F = 2,438; male: edf = 7.9, F = 2,959). Testosterone showed a marked sex divergence, with a steep rise in males (edf = 7.8, F = 7,743) and a more modest increase in females (edf = 5.7, F = 1,933). Estradiol, measured in females only, showed a nonlinear increase across the age range (edf = 5.4, F = 975).

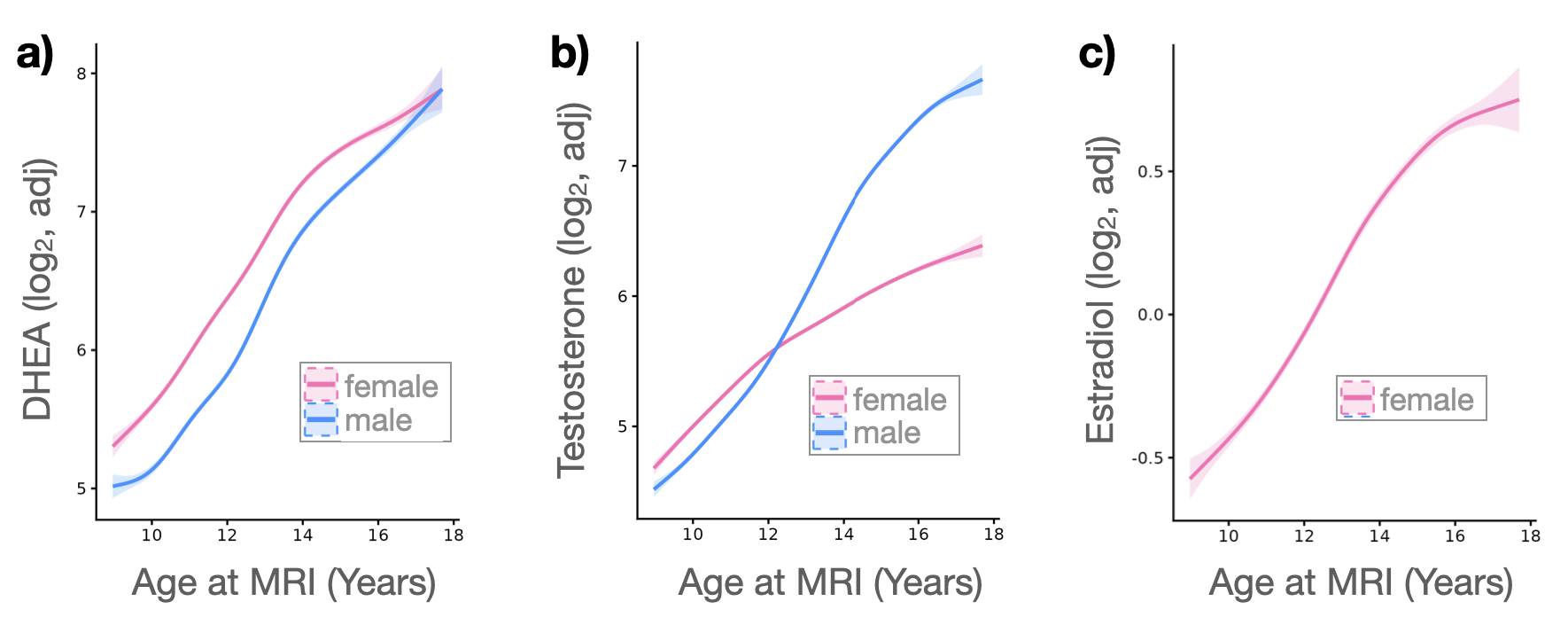

**Figure 5.** **Non-linear associations between age at MRI and hormone concentrations.**

Pubertal Development Scale (PDS) scores likewise followed strongly nonlinear sex-stratified age trajectories (Supplementary Fig. 6). Raw PDS scores, on aggregate, increased from approximately 1.5 near baseline to over 3 by age 17 in both sexes, with females advancing earlier (female: edf = 8.2, F = 10,338; male: edf = 8.2, F = 6,292). Sex-standardized (z-scored) PDS trajectories confirmed this sex difference, with female z-scores rising earlier and decreasing in velocity sooner than male z-scores (female: edf = 8.1, F = 8,382; male: edf = 8.3, F = 7,322). Two pseudo-qualitative observations are made here. First, self-report puberty development follows a more sigmoidal shape than hormones, that is reminiscent of that seen in pituitary dynamics. Second, because PDS questionnaires ultimately score on sex-specific items, their scales are challenging to harmonize, and thereby support sex stratified analyses.

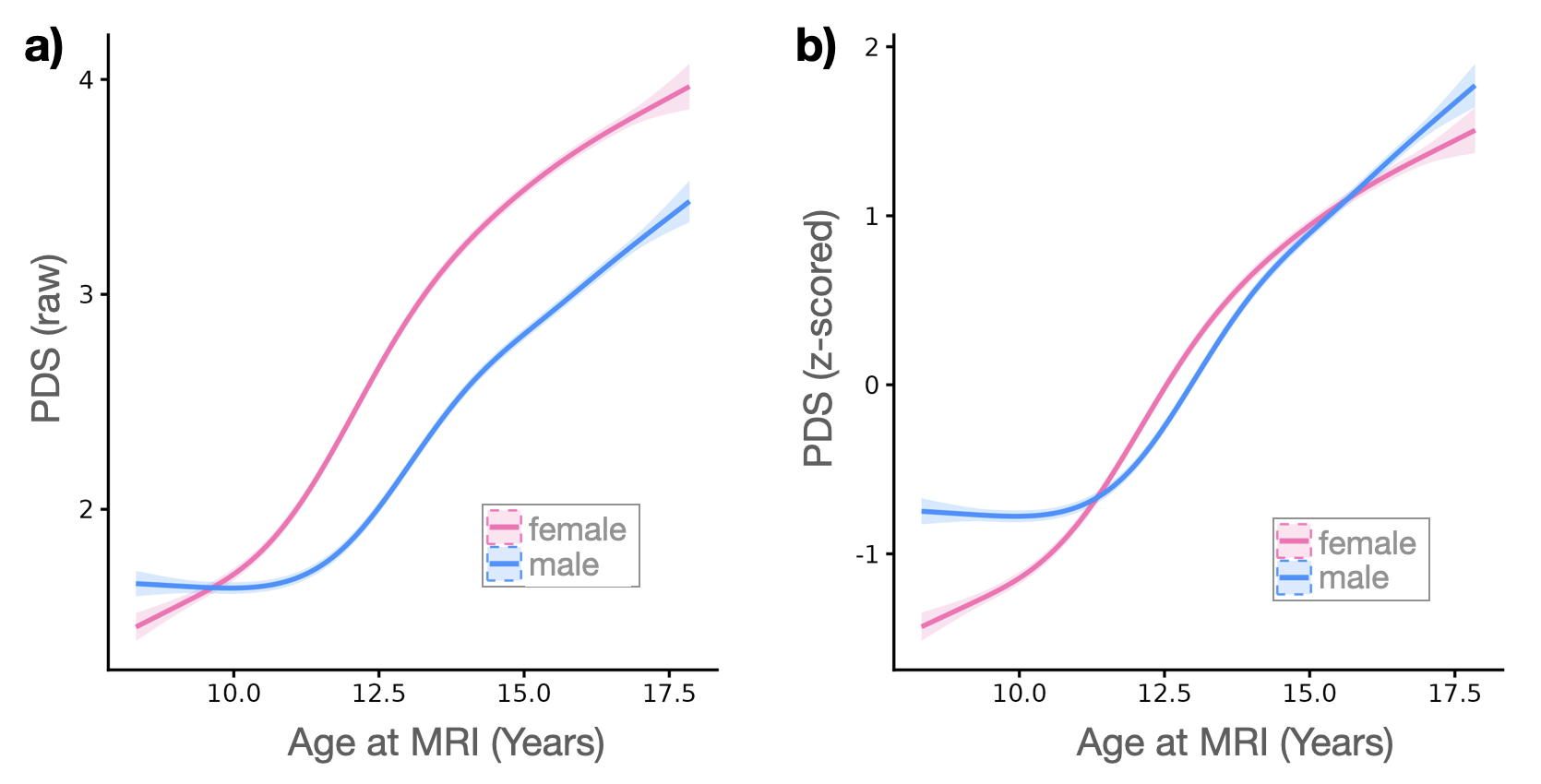

**Figure 6.** **Non-linear associations between age at MRI and PDS (raw and normalized).**

Pearson correlation matrices computed separately by sex (Supplementary Fig. 7) confirmed expected interrelationships among age, pubertal staging, hormones, and pituitary volume. In females, pituitary volume correlated most strongly with PDS (r = 0.69) and age (r = 0.66), with moderate correlations with DHEA (r = 0.49), testosterone (r = 0.46), and estradiol (r = 0.36). In males, pituitary volume correlated most strongly with age (r = 0.64), testosterone (r = 0.61), and PDS (r = 0.60). The higher pituitary–PDS correlation relative to any single hormone is consistent with PDS indexing a composite of pubertal processes rather than a single endocrine axis. Male estradiol was excluded from the correlation matrix as it was not analyzed in the main manuscript.

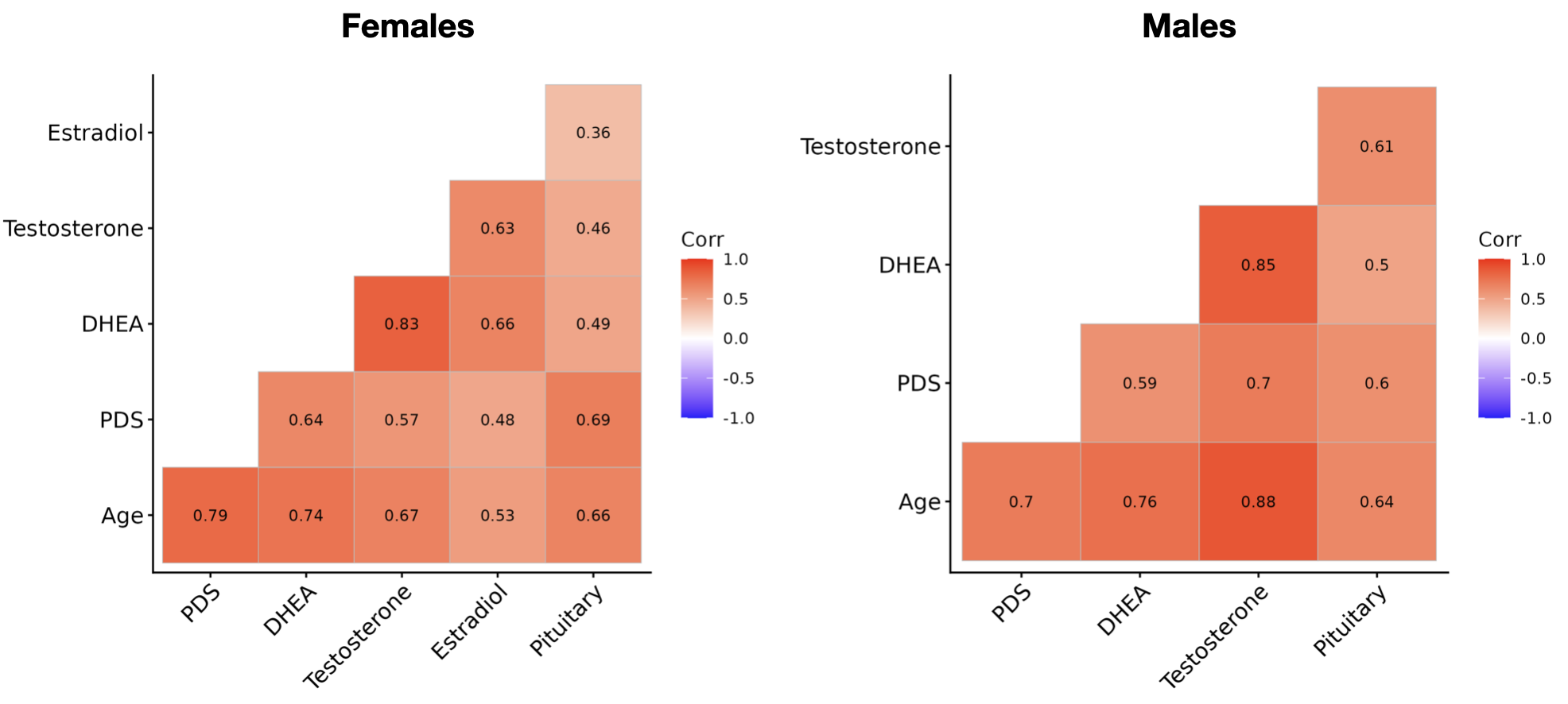

**Figure 7.** **Raw inter-relationships between key measures used in primary analysis.**
